# Novel biomarkers for glycaemic deterioration in type 2 diabetes: an IMI RHAPSODY study

**DOI:** 10.1101/2021.04.22.21255625

**Authors:** Roderick C Slieker, Louise A Donnelly, Livia Lopez-Noriega, Hermine Muniangi-Muhitu, Elina Akalestou, Mahsa Sheikh, Eleni Georgiadou, Giuseppe N. Giordano, Mikael Åkerlund, Emma Ahlqvist, Ashfaq Ali, Marko Barovic, Gerard A Bouland, Frédéric Burdet, Mickaël Canouil, Iulian Dragan, Petra JM Elders, Celine Fernandez, Andreas Festa, Hugo Fitipaldi, Phillippe Froguel, Valborg Gudmundsdottir, Vilmundur Gudnason, Mathias J. Gerl, Amber A van der Heijden, Lori L Jennings, Michael K. Hansen, Min Kim, Isabelle Leclerc, Christian Klose, Dmitry Kuznetsov, Dina Mansour Aly, Florence Mehl, Diana Marek, Olle Melander, Anne Niknejad, Filip Ottosson, Imre Pavo, Alexander Efanov, Kevin Duffin, Timothy J. Pullen, Kai Simons, Michele Solimena, Tommi Suvitaival, Asger Wretlind, Peter Rossing, Valeriya Lyssenko, Cristina Legido Quigley, Leif Groop, Bernard Thorens, Paul W Franks, Mark Ibberson, Joline WJ Beulens, Leen M ’t Hart, Ewan R Pearson, Guy A Rutter

## Abstract

We have deployed a multi-omics approach in large cohorts of patients with existing type 2 diabetes to identify biomarkers for disease progression across three molecular classes, metabolites, lipids and proteins. A Cox regression analysis for association with time to insulin requirement in 2,973 patients in the DCS, ANDIS and GoDARTS cohorts identified homocitrulline, isoleucine and 2-aminoadipic acid, as well as the bile acids glycocholic and taurocholic acids, as predictive of more rapid deterioration. Increased levels of eight triacylglycerol species, and lowered levels of the sphingomyelin SM 42:2;2 were also predictive of disease progression. Of ∼1,300 proteins examined in two cohorts, levels of GDF-15/MIC1, IL-18RA, CRELD1, NogoR, FAS, and ENPP7 were associated with faster progression, whilst SMAC/DIABLO, COTL1, SPOCK1 and HEMK2 predicted lower progression rates. Strikingly, identified proteins and lipids were also associated with diabetes incidence and prevalence in external replication cohorts. Implicating roles in disease compensation, NogoR/RTN4R improved glucose tolerance in high fat-fed mice and tended to improved insulin signalling in liver cells whilst IL-18R antagonised inflammatory IL-18 signalling towards nuclear factor kappa-B *in vitro*. Conversely, high NogoR levels led to islet cell apoptosis. This comprehensive, multi-disciplinary approach thus identifies novel biomarkers with potential prognostic utility, provides evidence for new disease mechanisms, and identifies potential therapeutic avenues to slow diabetes progression.

## INTRODUCTION

Type 2 diabetes is a progressive multifactorial disease which presently affects >400m worldwide, with numbers expected to increase to > 700 m by 2045.^1^ Biomarkers for the disease, which provide a deeper understanding of the disease process, are therefore eagerly sought. Importantly, their identification may improve prediction and personalized approaches to disease treatment.^2^

Whilst many studies have examined associations between circulating biomarkers and incident disease^3, 4^, to date few studies have explored changes associated with glycaemic deterioration after the development of diabetes. Published studies (reviewed in ^5^) have established that faster glycaemic deterioration is seen in those who are diagnosed younger, are more obese at diagnosis, have lower HDL, and higher HbA1c. A few studies have investigated genetic variants associated with more rapid progression with small and variable results^5^, although a report on a Hong Kong Chinese population reported a replicated finding that a high polygenic risk score consisting of 123 T2D risk variants was associated with increased progression to insulin requirement.^6^ To date, no studies that have adopted a multi-omic approach to biomarker discovery, or reported systematically how metabolites of different classes impact on progression. Such associations have the potential to be clinically useful in terms of prediction, as well as providing biological insights into the processes that drive glycaemic deterioration in T2D.

In a collaboration based around the EU Innovative Medicines Initiative-2 Risk Assessment and ProgreSsiOn of Diabetes (RHAPSODY) we have undertaken here to identify, in three large European cohorts, biomarkers of diabetes progression of three molecular classes: charged small molecules (metabolites), lipids and proteins. In this way, we identify novel species and, in the case of two of the identified proteins, provide evidence through functional studies in preclinical models for novel mechanisms of action in disease-relevant tissues.

## METHODS

### Discovery cohorts

Specific details on DCS^7^, GoDARTS^8^ and ANDIS^9^ have been described elsewhere.^10^ These cohorts were selected based in part on satisfactory quality control for biomarkers stability in stored samples.

Briefly, the Hoorn Diabetes Care System (DCS) cohort is a prospective cohort with currently over 14,000 individuals with routine care data. The Ethical Review Committee of the VU University Medical Center, Amsterdam approved the study. In 2008-2014, additional blood sampling was done in 5,500 participants, who provided written informed consent. These samples were used for this study. The turbidimetric inhibition immunoassay for haemolyzed whole EDTA blood (Cobas c501, Roche Diagnostics, Mannheim, Germany) was used to measure HbA1c. HDL (mmol/L) was measured enzymatically (Cobas c501, Roche Diagnostics). C-peptide was measured on a DiaSorin Liaison (DiaSorin, Saluggia, Italy).

The Genetics of Diabetes Audit and Research Tayside Study (GoDARTS) is a cohort of ∼8,000 patients with T2D. The study was approved by the Tayside Medical Ethics Committee and all individuals provided informed consent. Laboratory measurements were measured in a non-fasted state. C-peptide was measured on a DiaSorin Liaison (DiaSorin, Saluggia, Italy).

In the All New Diabetics in Scania (ANDIS) cohort, people with incident diabetes within Scania County, Sweden were recruited from January 2008 until November 2016 and all participants gave written informed consent. Regional ethics review committee in Lund approved the study. An electro-chemiluminescence immunoassay was used to measure C-peptide on a Cobas e411 (Roche Diagnostics, Mannheim, Germany) or a radioimmunoassay (Human C-peptide RIA; Linco, St Charles, MO, USA; or Peninsula Laboratories, Belmont, CA, USA). The Clinical Chemistry database was used to obtain HbA1c levels.

### Validation cohorts

External validation was performed in four external cohorts, ACCELERATE, AGES-Reykjavik, MDC-CC and DESIR. ACCELERATE is a clinical trial aimed at investigating the effect of evacetrapib on major adverse cardiovascular outcomes and has been described elsewhere.^11^ For the current study we only included the 6,054 individuals in the untreated arm. From this group, we selected 2,978 individuals with type 2 diabetes. In this group, 1003 individuals were excluded that did not have C-peptide levels or HbA1c levels, 72 were excluded because the age at diagnosis was < 35 years, 31 were excluded because they were on insulin at baseline and 22 were excluded because they had > 2 non-insulin glucose-lowering drugs and HbA1c levels > 8.5%. The final set consisted of 1,850 individuals of which 162 reached the primary endpoint.

AGES-Reykjavik is a prospective population-based study from Iceland.^12, 13^ In fasted blood samples protein levels were measured with the Somalogic platform. At baseline there were 4784 individuals free of diabetes and 654 with type 2 diabetes. Of 2,940 individuals free of diabetes at baseline and with 5-year follow-up information, 112 developed type 2 diabetes. ^13^ Identified proteins were tested against incident and prevalent type 2 diabetes using logistic regression, adjusted for age and sex.

Malmö Diet and Cancer Cardiovascular Cohort (MDC-CC) is a population-based cohort comprised of people living Malmö.^14^ Lipids were measured using the Lipotype platform in 3,667 individuals of which 555 developed type 2 diabetes.^15^ Proteins were measured using the Olink Proseek Multiplex proximity extension assay in 4915 individuals of which 700 developed type 2 diabetes. Identified lipids and proteins were tested against incident diabetes using Cox proportional hazard model adjusted for age, sex and BMI.

DESIR is a prospective population-based cohort comprised of middle-aged European individuals. Metabolomics was measured by Metabolon (Durham, NC).^16^ Logistic regression adjusted for age, sex and BMI was used to tested for an association between metabolites and prevalent (*n* = 43) and incident (*n* = 231) type 2 diabetes versus controls (*n* = 813).

### Molecular measurements

In the three discovery cohorts, those were selected with an age at diagnosis > 35 years, GAD negative, GWAS data and with a blood sample within three years after diagnosis. The metabolomics, lipidomics and proteomics groups were of different sizes (see further details below). Individuals were ranked based on the time between diagnosis and sampling date and those with the smallest time between diagnosis and sampling were selected. For metabolomics, we selected 1,267 in DCS, 900 in GoDARTS and 900 in ANDIS. For lipidomics, 900 individuals were selected in DCS, GoDARTS and ANDIS. For proteomics, we selected 600 individuals in DCS and GoDARTS.

### Small charged molecule analytes

In 2,973 individuals 19 small charged molecule analytes (referred to as metabolomics) using UHLPC-MS/MS (UHLPLC: 1290 Infinity system from Agilent Technologies, Santa Clara, CA, USA; MS/MS: 6460 triple quadrupole system from Agilent Technologies) relevant to diabetes were quantified, which was used as the discovery set.^17^ In DCS, all samples passed QC and were used in the analysis. In GoDARTS, three failed in QC and the remaining samples were used for analysis. In ANDIS, 4 failed QC and of the 892 remaining samples, 811 were free of the outcome at sampling. In addition, a validation set was generated comprised of 2,668 individuals (699 GoDARTS, 1,969 ANDIS).

### Lipid measurements

Six hundred and fourteen (614) lipids were determined using a QExactive mass spectrometer (Thermo Scientific) equipped with a TriVersa NanoMate ion source (Advion Biosciences) on the Lipotype lipidomics platform (Lipotype, Dresden, Germany).^18^ Samples (*n* = 2,608) were measured in batches of 84 samples each. Unprocessed mass spectra files were used to identify lipids with LipotypeXplorer.^19^ Lipid identifications with a signal-to-noise ratio > 5, and a signal intensity 5-fold higher than in corresponding blank samples were considered for further data analysis. Eight reference samples were used to apply batch correction and amounts were further adjusted for analytical drift (p-value slope ≤ 0.05 and R^2^ ≥ 0.75 and the relative drift > 5%). Lipid nomenclature is used as described previously and SwissLipids database identifiers are provided (**Table S1**).^20^ After quality control 162 lipid species were used in this study. The median coefficient of subspecies variation of the 162 lipids used as accessed by reference samples was 9.49% across all three cohorts. In DCS, 900 individuals were included for lipidomics measurements, all passed QC, and all were suitable for analysis. In GoDARTS, 898 individuals were included in the analysis, 1 failed QC and all 897 remaining samples were included in the analysis. In ANDIS, 896 individuals were included in the analysis, 5 failed QC and of the 891 remaining samples, 811 were free of the outcome at sampling.

### Protein measurements

Proteins were measured on the SomaScan® Platform from Somalogic (*n* = 1195 proteins) on the SomaLogic SOMAscan platform (Boulder, Colorado, USA) in 1,188 individuals. Top associated proteins were validated in ANDIS (*n* = 1992) and ACCELERATE (*n* = 1850) using ELISA with time to insulin requirement as the outcome. External validation was performed for the top proteins based on P-value and/or effect size for which an ELISA was available or could be developed.

### Primary endpoint

The primary endpoint time to insulin requirement was defined as the period from diagnosis to a clinical endpoint of the earlier of (i) starting sustained (more than 6 months duration) insulin treatment or (ii) clinical requirement of insulin as indicated by two or more HbA1c measurements>8.5% more than three months apart when on two or more non-insulin diabetes therapies.^21^ In DCS, 600 individuals were included for proteomics measurements, 11 failed QC and all were included for analysis. In GoDARTS, 600 individuals were included in the analysis, 1 failed QC and the 599 remaining samples were included in the analysis.

### Federated database

All main analyses were performed on a federated database system. Opal, an open-source data warehouse (Open Source Software for BioBanks, OBiBa) was used to store cohort data on local nodes and remote analysis was performed in R using *DataSHIELD*^22^ and *dsSwissKnife* R packages.^23^ A central server was set up at the Swiss Institute of Bioinformatics to manage federated node access, user administrator and software deployment. Local nodes were set up at the respective cohorts. All data was harmonized according to the CDISC Study Data Tabulation Model (www.cdisc.org) prior to inclusion into the federated database.

### Mendelian randomisation

Genetic instruments for proteins, lipids and metabolites predictive of progression in the three cohorts were obtained from published GWASs. Protein quantitative trait loci (QTLs) were obtained from Gudmundsdottir et al.^13^ Lipid QTLs were obtained from Tabassum et al.^24^ Metabolite QTLs were obtained from Lotta et al.^25^ Only QTLs with P-values < 5·10^-8^ were included. For traits with one instrument Wald ratio was used and for multiple instruments inverse variance weighting. Instruments were excluded when in LD (r^2^ > 0.1). Genetic instruments for type 2 diabetes were obtained from the latest GWAS on incident type 2 diabetes.^26^ Horizontal pleiotropy was estimated based on MR-Egger intercept. Cochran Q-statistic was used to estimate heterogeneity of instruments.

### Cells and cell culture

HEK-Blue IL-18 cells passages 1-16 (InvivoGen, USA) were cultured in 4.5 g/L glucose Dulbecco’s Modified Eagle’s Medium (DMEM) supplemented with 10% foetal bovine serum (FBS), 2 mM L-Glutamine, 50 U/ml penicillin, 50 μg/ml streptomycin (Sigma, UK) and 100 μg/ml Normocin (InvivoGen, USA). HEK-Blue IL-18 cells were designed to detect bioactive IL-18 by monitoring the activation of the NF-κB and AP-1 pathways. They were generated by stable transfection of HEK293 derived cells with the genes encoding IL-18R and IL-18 receptor accessory protein (IL-18RAP). Additionally, the TNF-α and the IL-1β responses have been blocked to guarantee a specific respond to IL-18. Cells were seeded in T75 flasks and sustained at 37°C in a humidified incubator containing 5% CO2. Experiments were carried out with HEK293 cells as well as a negative control.

### Cell transfection

HEK-Blue IL-18 and HEK293 cells were seeded in 12-well plates at 30,000 per well and transfected after 48 hours with both NF-κB and Renilla using Lipofectamine 2000 DNA Transfection Reagent (Invitrogen, USA). For each well, 1.5 μl of Lipofectamine reagent was diluted in 100μl Opti-MEM Gibco medium (Thermo Fisher, USA). The amount of 1 μl of NF-κB DNA at a concentration of 472.6 ng/μl and 1 μl of Renilla at a concentration of 40 ng/μl were also diluted in 100 μl of Opti-MEM medium for each well, the diluted DNA mix was then added to diluted Lipofectamine reagent and the transfection mix was incubated for 20 minutes at RT. Cells were washed with 1ml of PBS and 400 μl of Opti-MEM medium was added per well. Cells were transfected with 200 μl transfection mix each well. Medium was then changed to complete growth medium (DMEM supplemented with 10% FBS, 2 mM L-Glutamine, 50 U/ml penicillin, 50 μg/ml streptomycin and 100 μg/ml Normocin) after 4 hours.

### Cell treatment and stimulation

Transfected cells were stimulated with different concentrations of IL-18, IL-18Rα or both to measure changes in NF-κB activation. Concentrations were made by diluting different volumes of 1 μM IL-18 and IL-18Rα stock (diluted from freeze-dried powder in in distilled water) in serum-free medium to exclude the effect of serum factors and cells were stimulated with 300 μl of each condition per well of 12-well plate for 6 hours. Recombinant human IL-18 and rhIL-18 Rα/Fc chimera were obtained from MBL and R&D Systems respectively. Normal rabbit IgG from Abcam (ab171870) was used as a negative control.

### Dual luciferase reporter assay

Cells transfected with NF-κB and stimulated with IL-18/IL-18Rα were lysed following six hours of treatment by adding 200 μl of Promega Passive Lysis Buffer (PLB) to each well and gentle shaking for 15 minutes at RT. Cell lysates were stored at -20°C and luciferase activity was measured the next day using Promega Dual-Luciferase Reporter Assay System according to manufacturer’s instructions to study gene expression at the transcriptional level. Briefly, Luciferase assay reagent II (LAR II) was dispensed into a luminometer tube for each condition. Cell lysate was resuspended in tube containing LAR II and Firefly luciferase activity was measured using Berthold Lumat LB 9507 Tube Luminometer. Next, Stop & Glo reagent was added to each tube and the Renilla luciferase activities were measured. Measurements were read with PuTTY software and all Firefly luciferase activities were normalized with Renilla luciferase activities to obtain the NF-κB signalling ratios using Microsoft Excel software.

### HepG2 cells stimulation and Western (immuno-) blotting

HepG2 Cells (human hepatocytes) were cultivated in DMEM media with 1g/L of glucose, supplemented with 10% SVF. HepG2 cells line were maintained at 37 °C and 5% CO2. Cells were treated with 0, 1nM, 10 nM, or 100 nM of NogoR (see above) or CRELD1 (Mouse Fc-tagged, Sino Biological, Cat # 51149-M02H) recombinant proteins for 3 hours and then stimulated with insulin for 15 minutes prior protein extraction. Cells were lysed in radioimmunoprecipitation assay buffer (RIPA buffer: 20 mM Tris-HCl, 50 mM NaCl, 1 mM Na_2_-EDTA, 1 mM EGTA, 1% NP-40, 1% sodium deoxycholate) with 1% Phosphatase Inhibitor Cocktails (P0044 and P5725, Sigma-Aldrich) and 1% Protease Inhibitor Cocktail (P8340, Sigma-Aldrich). Protein concentration was measured using a Pierce BCA Protein Assay Kit (Thermofisher, 23225) at 562 nm using a PHERAstar reader (BMG Labtech). After protein transfer to a polyvinylidene Difluoride (PVDF) membrane (Millipore), membranes were blocked with Tris-buffered saline 1X plus 0.1% (v/v/) Tween (TBST), containing 4% (w/v) BSA for 1h. Primary antibody incubation was performed overnight during rotation at 4°C in TBST-4% (w/v) BSA and horseradish peroxidase (HRP)-conjugated secondary antibody incubation was subsequently performed for 1 h in TBST-4% (w/v) milk powder. For development we used Clarity Western ECL Substrate (Bio-Rad).

### In vivo metabolic tests

All *in vivo* procedures were approved by the UK Home Office, according to the Animals (Scientific Procedures) Act 1986 with local ethical committee (Imperial AWERB) approval under personal project license (PPL) number PA03F7F07 to I.L. For the oral glucose tolerance test (OGTT), C56BL6J mice (Charles River) maintained for four weeks on a high fat high sucrose (HFHS) diet (a 58 kcal% Fat and Sucrose diet (D12331, Research Diet, New Brunswick, NJ) were fasted for 16 h prior to experiment and received an oral glucose load (2 g/kg of body weight). For insulin tolerance tests, mice received an intraperitoneal injection of insulin (1 IU/kg) after 3 h fasting. Blood glucose levels were determined by tail venepuncture using a glucose meter (Accu-Chek; Roche, Burgess Hill, UK) at 0, 15, 30, 60 and 120 min. after the glucose load. For insulin measurements blood was collected in EDTA covered tubes at times 0, 15 and 30 min. after the glucose load (2 g/kg of body weight). Subsequently, blood was centrifuged at 4000 x g for 20 min. at 4 °C and plasma was collected. Insulin was determined by ELISA (CrystalChem, 90080), according to the manufacturer’s instructions. Daily intraperitoneal injections of NogoR were performed with indicated dose of NogoR (mouse, His and Fc tag; Sino Biologicals Cat # 50106-M03H).

### Insulin secretion from mouse and human islets

Mouse pancreatic islets were isolated from male C57BL/6 mice (Envigo, Indianapolis, IN) by collagenase digestion. Use of animals was approved by Eli Lilly and Company’s Institutional Animal Care and Use Committee. Human pancreatic islets from listed cadaver organ donors that were refused for pancreas or islet transplantation were obtained from Prodo Labs (Irvine, CA) and InSphero AG (Schlieren, Switzerland) and were used in accordance with internal review board ethical guidelines for use of human tissue. Islets were cultured in the complete PIM(S) Prodo Islet Media (Prodo Labs) and RPMI 1640 medium (Invitrogen) supplemented with 11 mm glucose, 10% (v/v) heat-inactivated foetal bovine serum (Invitrogen), 100 IU/ml penicillin, and 100 μg/ml streptomycin (Invitrogen).

For insulin secretion in mouse islets, islets were incubated for 30 min. in Earle’s balanced salt solution (EBSS) buffer supplemented with 3 mM glucose and 0.1% BSA. Then groups of three islets were selected and cultured with tested proteins at indicated glucose concentration in 300 µL of EBSS for 60 minutes at 37°C. At the end of incubation, the supernatant was collected and subjected to insulin analysis. To measure insulin secretion in human pancreatic islets, single islets placed in a GravityTRAP 96-well plate (InSphero) were washed and incubated for 30 minutes in 100 µL of EBSS supplemented with 0.1% BSA and 3 mM glucose. Then the buffer was replaced with 100 µL of EBSS containing indicated glucose and protein concentrations and further cultured for 60 minutes at 37°C. At the end of incubation, the supernatant was collected and submitted for insulin analysis. Insulin levels were determined with the Meso Scale Discovery (Gaithersburg, MD) electrochemiluminescence insulin assay.

For chronic incubation experiments, after overnight recovery human or mouse islets were cultured in 12 well plate (20-30 islets per well) in the RPMI-1640 culture media containing tested proteins for 72 h. At the end of incubation, islets were transferred into EBSS supplemented with 3mM glucose and 0.1% BSA. Then, 1h insulin secretion in response to elevated glucose in islets pre-treated with proteins was measured.

### Quantification of β-cell proliferation

After overnight recovery, mouse or human islets were cultured for 72h in 12-well plates (200-300 islets per well) in RPMI-1640 medium containing 5mM glucose, 2% FBS, 10 µM EdU and tested proteins. At the end of incubation, islets were washed, dispersed into single cells with the Accutase solution (Sigma) and placed in 96 well plate coated the Cell-Tak Cell and Tissue Adhesive (Corning). Cells were fixed, permeabilized and stained with Click-iT EdU HCS assay (ThermoFisher), PDX1 antibody (ab47308, Abcam) and Hoechst 33342 (ThermoFisher) nucleic acid dye. Cell images were captured and analysed using InSight Imaging System (ThermoFisher).

### Islet cell apoptosis

After 3-4 day culture, mouse or human islets were plated (1 islet per well) in the GravityTRAP 96-well plate (InSphero) in 100 µl/well RPMI-1640 medium containing 11 mM glucose and 1% FBS. To induce cell death, islets were treated with glucose (25 mM) and palmitic acid (300 µM palmitate conjugated with BSA), cytokine mixture (120 ng/ml TNF-α, 60 ng/ml IL-1B and 240 ng/ml IFN-γ, R&D Systems) or tunicamycin (0.03 µg/ml, Tocris). After 72-hour incubation, islets were caspase activity was measured with the Caspase-Glo 3/7 Assay (Promega) according to manufacturer’s protocol.

### Statistical analyses

A Cox proportional hazard model was used to identify molecular risk factors for time to insulin requirement in R (v3.6.0) remotely on each cohort federated node using the *dssCoxph* function in *dsSwissKnife*. Three models with and without biomarkers were explored, where a primary model was adjusted for age, sex and BMI. A second model was further adjusted for HDL and C-peptide and the fully adjusted model additionally for diabetes duration and the use of glucose lowering drugs. Models were stratified for HbA1c (strata: < 53 mmol/mol, 53-64 mmol/mol and > 64 mmol/mol). Figures and meta-analysis was performed locally with R (v4.0.3). Estimates in each cohort were meta-analysed using the *metagen* function from the R package meta. P-values were adjusted for multiple testing using the Benjamini-Hochberg procedure and a P_FDR_ < 0.05 was considered significant. Figures were made using ggplot2 (v3.3.2). Analysis of cellular and metabolic data were performed using GraphPad Prism versions 7.0-9.0 (San Diego, CA, U.S.A.) deploying ANOVA with appropriate post hoc correction for multiple testing.

## RESULTS

### Cohort characteristics and modelling of glycaemic deterioration

Individuals from three cohorts, DCS, GoDARTS and ANDIS were included. In a subset molecular characterization was performed of which characteristics are shown in **Table S2**. The characteristics across the cohorts were comparable (**Table S2**). Male subjects were more abundant in the cohorts (>55%), and the average age ranged from 61-67 years with a BMI of 30-32 kg/m^2^. Glycated haemoglobin (HbA1c) levels were on average lowest in DCS (median[Q25-Q75]: 47.08[42-50] mmol/mol), followed by GoDARTS (55.54[46-61] mmol/mol) and ANDIS (60.06[46.0-68.0]). The time from diagnosis to sampling time ranged from 0 to 2.63 years. Three phenotypic models were explored in the included cohorts which showed concordance with BMI, use of glucose-lowering drugs being risk factors and age, HDL and C-peptide being protective (**Table S3**).

### Metabolites are associated with increased diabetes risk and progression

Out of the 19 small metabolites examined, five were associated with disease progression with nominal significance in the base model (age, sex, BMI adjusted, P<0.05) in the meta-analysis of three cohorts. These were homocitrulline (Hcit), aminoadipic acid (AADA), isoleucine (Ile), glycocholic acid (GCA), taurocholic acid (TCA). Out of the five, the association of two remained significant after multiple testing adjustment, including aminoadipic acid (AADA, HR[CI]=1.11[1.04-1.19], P_FDR_ = 0.03) and homocitrulline (Hcit, 1.12[1.04-1.21], P_FDR_ = 0.04, **Fig. 1, Table S4).** Both metabolites showed associations in the same direction in the replication cohorts, but non-significant with attenuated effect sizes (AADA, HR[CI]=1.03[0.96-1.11]; Hcit 1.03[0.88-1.21]). In external validation cohorts, Hcit showed a trend as a risk factor for incident diabetes (HR[CI] = 1.05[0.74-1.48]) in MDC. Based on a logistic model in DESIR, Hcit was a risk factor for prevalent diabetes (OR[CI] = 1.32[1.05-1.66]), but not incident diabetes (HR[CI] = 0.97[0.73-1.30]). AADA has previously been associated with a higher risk of incident type 2 diabetes in Wang et al. (OR[CI] = 1.60[1.19-2.16]).^27^ Finally, the most consistent risk factor for time to insulin was isoleucine level, which was nominally significant in the discovery cohort (HR[CI] = 1.09[1.00-1.20]), a risk factor for incident diabetes in MDC (HR[CI] = 1.48[1.26-1.74]) and DESIR (OR[CI] = 23.88[3.13-182.31) as well as prevalent diabetes (OR[CI] = 10.94[3.94,30.32]). Finally, GCA and TCA were modest risk factors for time to insulin requirement, with hazard ratios of 1.09[1.01-1.17] and 1.06[1.01-1.12], respectively. In the replication set both TCA and GCA were in the same direction, but no longer significant with hazard ratios of 1.09[0.91-1.31] and 1.04[0.94,1.12].

**Figure 1.**
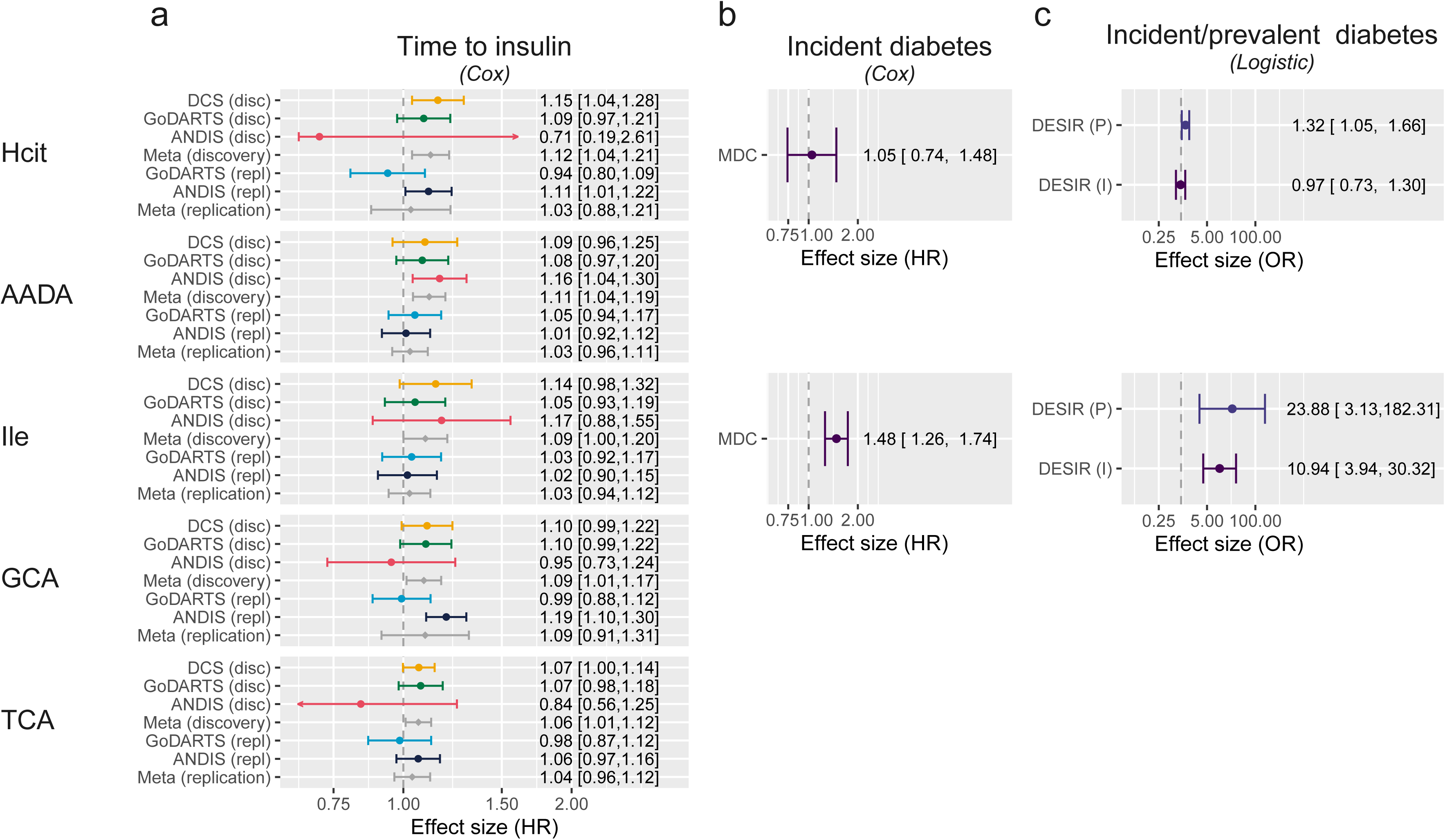
**Metabolites associated with diabetes development and progression. a)** Hazards of a time to insulin model in the three discovery cohorts plus two replication sets in two of the three cohorts and their respective meta-analyses (Model 1). The figure shows the five nominally significant metabolites, with Hcit and AADA being also significant after multiple testing. **b)** Hazards of incident diabetes in MDC based on a Cox proportional hazards model adjusted for age, sex and BMI. **c)** Odds ratios of incident and prevalent diabetes in DESIR based on a logistic regression model adjusted for age, sex and BMI.

### Plasma triglyceride levels are markers of diabetes progression and incident diabetes

Among the 162 lipids investigated, the levels of nine reached significance in the base model. Among these eight lipids were a risk factor for early insulin requirement, and these were all triglycerides (**Fig. 2, Table S5**). These eight lipids were also a risk factor for incident diabetes in MDC (**Fig. 2**). A single lipid was *protective* for early insulin initiation (SM 42:2;2, HR[CI] = 0.85[0.77-0.94]). Interestingly, SM 42:2;2 was a *risk factor* for incident diabetes in MDC (HR[CI] = 1.16[1.06-1.27]). Further adjustment in the discovery cohort did attenuate the effect size but the direction remained the same and four and three remained significant in the partly and fully adjusted model, respectively **(Table S5)**.

**Figure 2.**
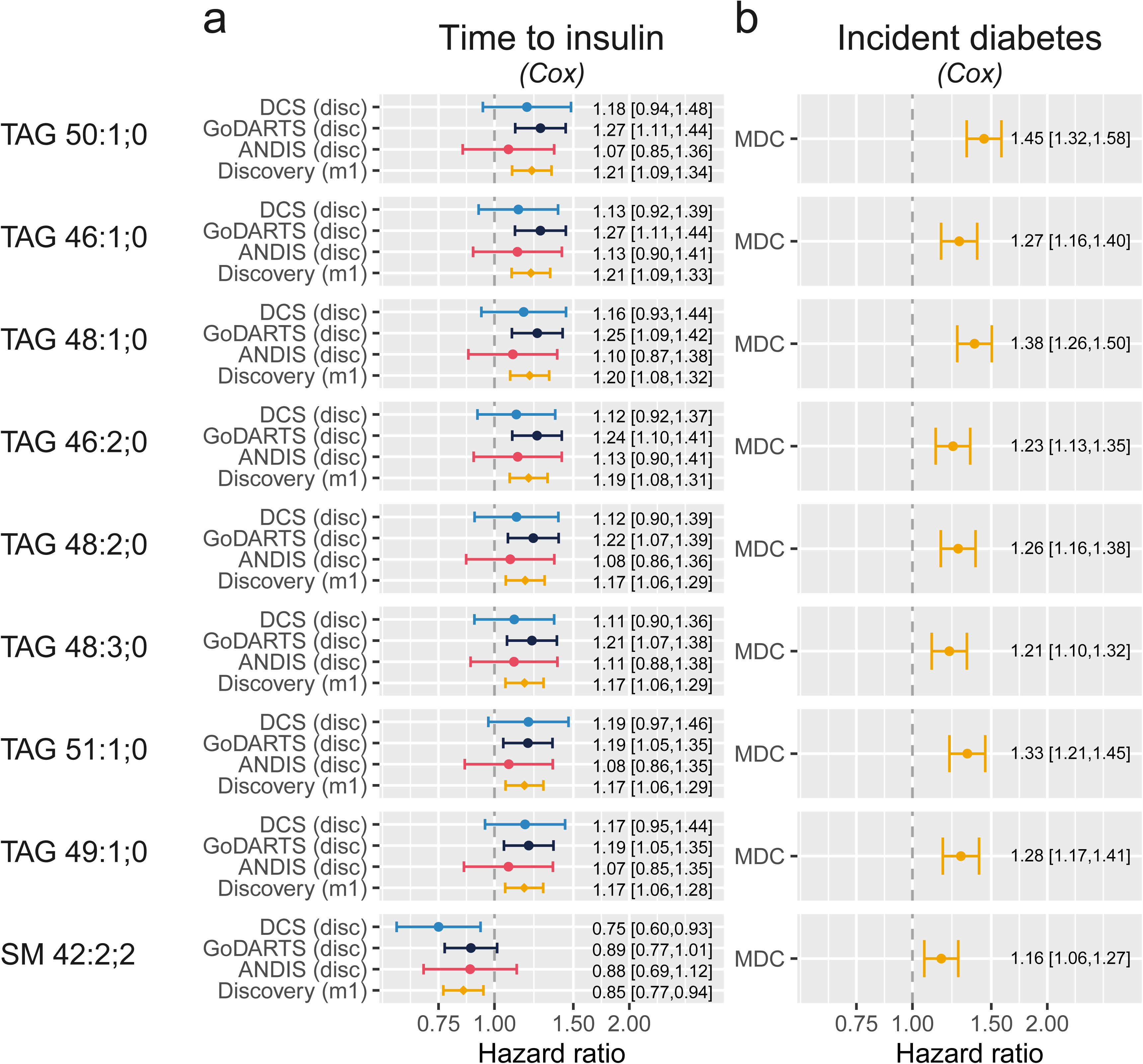
**Lipids associated with diabetes development and progression. a)** Hazards of a time to insulin model in the three discovery cohorts and the meta-analysed hazards (Model 1). The figure shows the nine significant lipids after multiple testing. **b)** Hazard models of incident diabetes in MDC based on a Cox proportional hazards model.

### Plasma proteins levels associate with diabetes progression and prevalent and incident diabetes

In the 1195 investigated plasma proteins, the levels of 98 were nominally associated with time to insulin in the base model. Additional adjustment attenuated the hazard ratios only minimally in both the partly and fully adjusted model. MIC-1/GDF15 was the protein associated with the highest risk of progression (HR[CI] = 1.34[1.17-1.54]) and this association was replicated in ACCELERATE (HR[CI] = 1.22[1.04-1.42]). The protein associated with the second highest risk of progression was the Nogo receptor (NogoR, HR[CI] = 1.33[1.03-1.72], **Fig. 3, Table S6**). In ANDIS, NogoR also replicated (HR[CI] = 1.20[1.07-1.34], **Fig. 3**). NogoR was also a risk factor for incident (OR[CI] = 1.45[1.15-1.83]) and prevalent diabetes in AGES-Reykjavik (OR[CI] = 1.77[1.60-1.95]). In the top associated proteins, four were protective including SMAC, coactosin-like protein, testican-1 and HEMK2, of which HEMK2 was the most protective (HR[CI] = 0.78[0.68-0.89]). In the AGES-Reykjavik study, HEMK2 was also protective for prevalent diabetes (OR[CI] = 0.78[0.72,0.85]).

**Figure 3.**
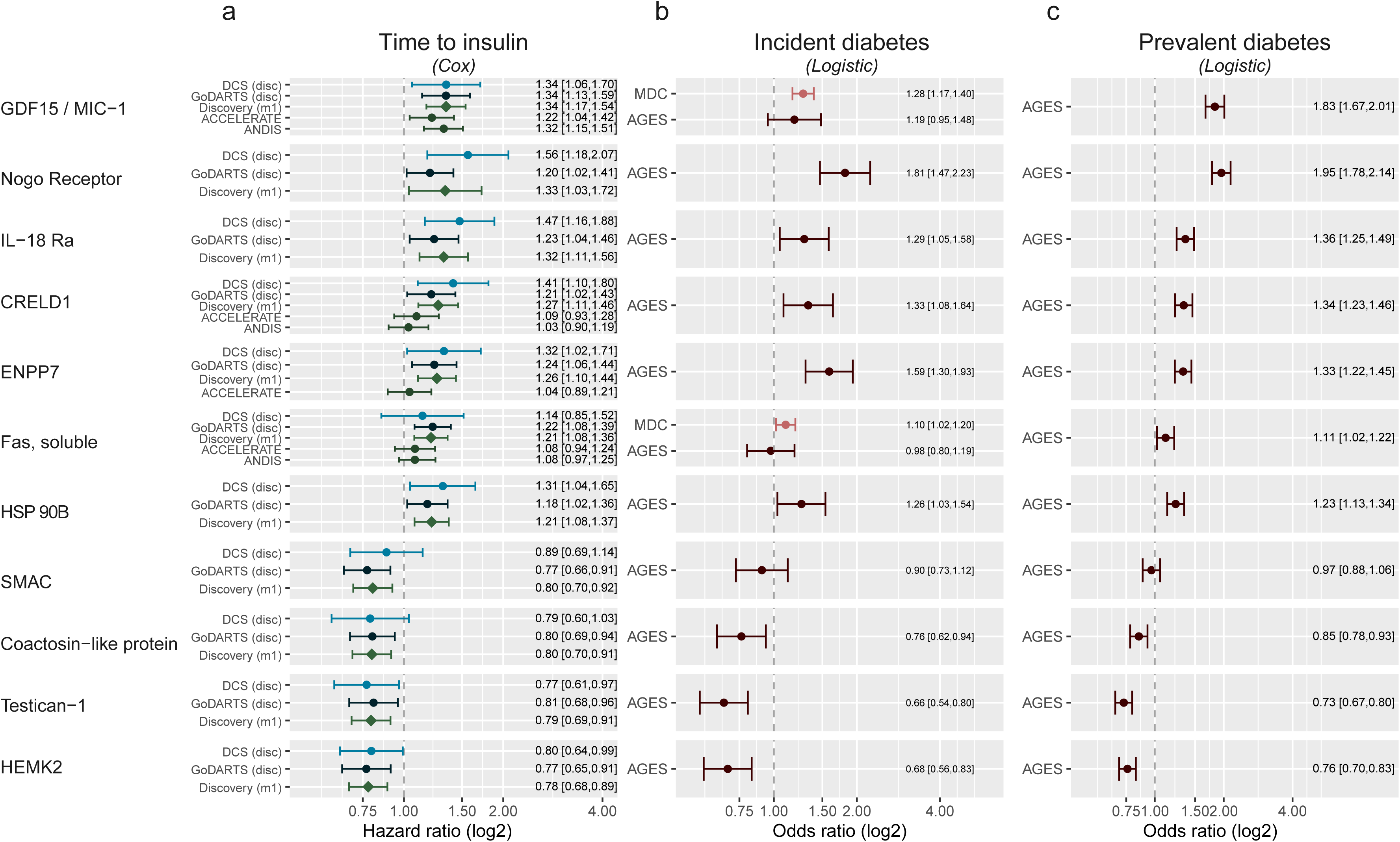
**Proteins in plasma or serum associated with time to insulin requirement. a)** Top proteins associated with time to insulin requirement. Shown is the top 10 based on P-value plus Nogo receptor, which showed the largest risk of the top hundred proteins. X-axis, hazard ratio on a log2 scale and studies on the y-axis. **b)** Association between protein levels and incident diabetes. X-axis, odds ratio on a log2 scale. **c)** Association between protein levels and prevalent diabetes. X-axis, odds ratio on a log2 scale.

### Evidence of causality of biomarkers on incident diabetes based on Mendelian Randomisation

To assess causality of the identified biomarkers we would ideally have tested against the genetics of time to insulin requirement in people with diabetes, but in the current study the outcome was underpowered (*n =*14000) and there is no publicly available data for time to insulin genetic variants. Instead, we investigated the causality of biomarkers on type 2 diabetes. We found no significant associations with incident diabetes for any of the top metabolites (**Table 1)**. However one of the nominally significant phosphatidyl ethanaolamines, PE 18:0;0-18:2;0 has support for a causal association with type 2 diabetes (**Table 1**, β[95%CI] = -0.07[-0.12 – -0.02], P = 3.89·10^-3^). For the proteins, modest evidence of a causal relation was observer for three proteins, GDF15 (β[95%CI] = 0.03[0.01– 0.05], P= 2.68·10^-3^), IL-18Ra (β[95%CI] = 0.02[0.003– 0.03], P= 0.014) and FAS (β[95%CI] = 0.05[0.005-0.09], P = 0.03). For the other protein biomarkers there was no evidence of a causal relation.

**Table 1.** Mendelian randomization analysis on metabolites, lipids and proteins against incident type 2 diabetes

Metabolites
| *variable* | *method* | *nsnp* | *Beta* | *Lower* | *Upper* | *P-value* | *Egger intercept* | *Q* | *Q(df)* | *Q(Pvalue)* |
| --- | --- | --- | --- | --- | --- | --- | --- | --- | --- | --- |
| AADA | IVW | 2 | 0.02 | -0.09 | 0.13 | 0.77 |  | 0.02 | 1 | 0.88 |
| Citrulline | IVW | 6 | -0.01 | -0.29 | 0.28 | 0.97 | -0.04 | 83.02 | 5 | <0.001 |
| Isoleucine | IVW | 5 | -0.06 | -0.94 | 0.83 | 0.90 | -0.11 | 127.69 | 4 | <0.001 |
| Leucine | IVW | 6 | -0.08 | -0.74 | 0.58 | 0.81 | -0.07 | 127.77 | 5 | <0.001 |

Lipids
| *variable* | *method* | *nsnp* | *Beta* | *Lower* | *Upper* | *P-value* | *Egger intercept* | *Q* | *Q(df)* | *Q(Pvalue)* |
| --- | --- | --- | --- | --- | --- | --- | --- | --- | --- | --- |
| PE 18:0;0_18:2;0 | IVW | 3 | -0.07 | -0.12 | -0.02 | $3.89 \cdot 10^{-3}$ | 0.02 | 5.53 | 2 | 0.06 |
| SM 42:2;2 | IVW | 2 | 0.00 | -0.07 | 0.07 | 0.99 |  | 0.91 | 1 | 0.34 |
| TAG 50:1;0 | IVW | 2 | 0.01 | -0.04 | 0.05 | 0.75 |  | 0.00 | 1 | 0.97 |

Proteins
| *variable* | *method* | *nsnp* | *Beta* | *Lower* | *Upper* | *pval* | *Egger intercept* | *Q* | *Q(df)* | *Q(Pvalue)* |
| --- | --- | --- | --- | --- | --- | --- | --- | --- | --- | --- |
| GDF15 | IVW | 8 | 0.03 | 0.01 | 0.05 | $2.68 \cdot 10^{-3}$ | 0.01 | 8.83 | 7 | 0.27 |
| IL-18Ra | IVW | 14 | 0.02 | 0.003 | 0.03 | 0.01 | 0.00 | 6.75 | 13 | 0.91 |
| FAS | IVW | 4 | 0.05 | 0.005 | 0.09 | 0.03 | 0.00 | 6.34 | 3 | 0.10 |
| RTN4R | IVW | 10 | -0.01 | -0.03 | 0.01 | 0.28 | -0.01 | 4.16 | 9 | 0.90 |
| ENPP7 | IVW | 4 | 0.01 | -0.01 | 0.02 | 0.34 | 0.00 | 0.85 | 3 | 0.84 |
| CRELD1 | IVW | 7 | 0.01 | -0.01 | 0.02 | 0.43 | 0.01 | 7.33 | 6 | 0.29 |

IVW, inverse variance weighting; nsnp, number of instruments

### Functional analyses of identified protein biomarkers

#### Glucose-stimulated insulin secretion

We chose to study six protein biomarkers with greatest effect size (GDF15, IL-18Ra, NogoR, CRELD1, FAS, ENPP7) and which accelerated progression, *i.e.*, those which may plausibly exert a deleterious effect on insulin secretion or action. None of these affected basal (3 mM glucose) or high (17 mM) glucose-stimulated insulin secretion (GSIS) acutely (1h) or after longer incubations (48h) from either mouse (**Fig. 4a,b**) or human (**Fig. 4c**) islets. Glucagon-like peptide-1 (GLP-1) was used as a positive control, and stimulated secretion at the higher glucose concentration, as expected.

**Figure 4.**
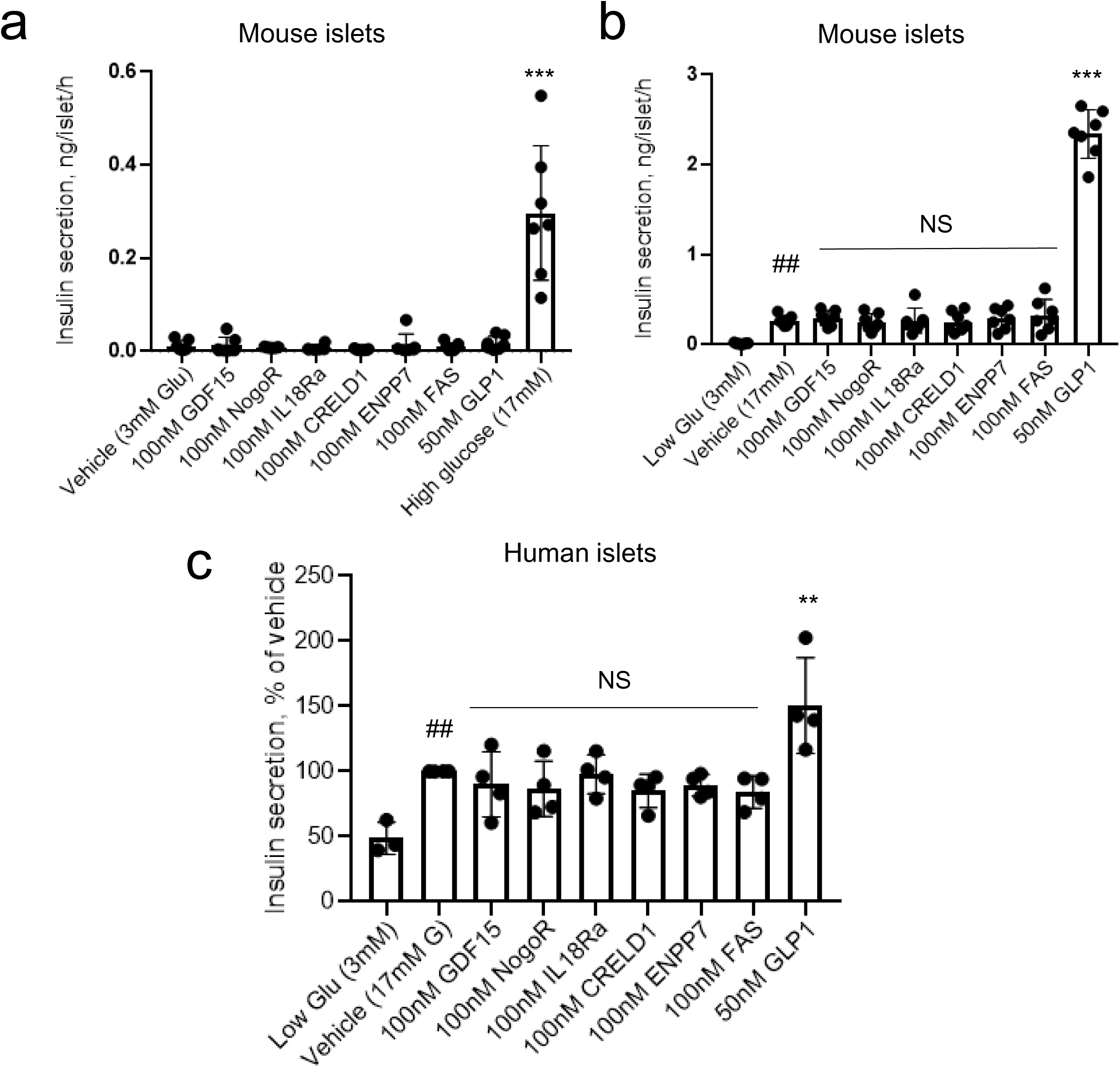
**Impact of identified biomarkers on insulin secretion from mouse (a,b) and human** (c) islets. Incubations were performed for 30 min. at the indicated concentrations of glucose, and secreted insulin measured using an electrochemiluminescence assay. ***, ** p = 0.001, 0.01 compared to vehicle; ##p<0.01 vs 3 mM glucose. Comparisons by one-way ANOVA in each case. Other details are given in the Methods Section.

To determine whether any of the examined protein biomarkers might affect pancreatic beta cell mass, we next assessed their impact on beta cell apoptosis (**Fig. 5a-c**) and on proliferation (d) in mouse islets. Of those examined, only IL-18Ra (3-fold), and NogoR (> 15-fold) exerted an effect, increasing apoptosis in mouse islets. Dose response analyses (**Fig. 5b**) revealed that the effects of NogoR were apparent only at high concentrations (> 1 nM) likely to be above the normal physiological range. At 100 nM NogoR, but not IL-18Ra also enhanced apoptosis in human islets (**Fig. 5c**). None of the tested compounds affected human islet proliferation (**Fig. 5d**)

**Figure. 5.**
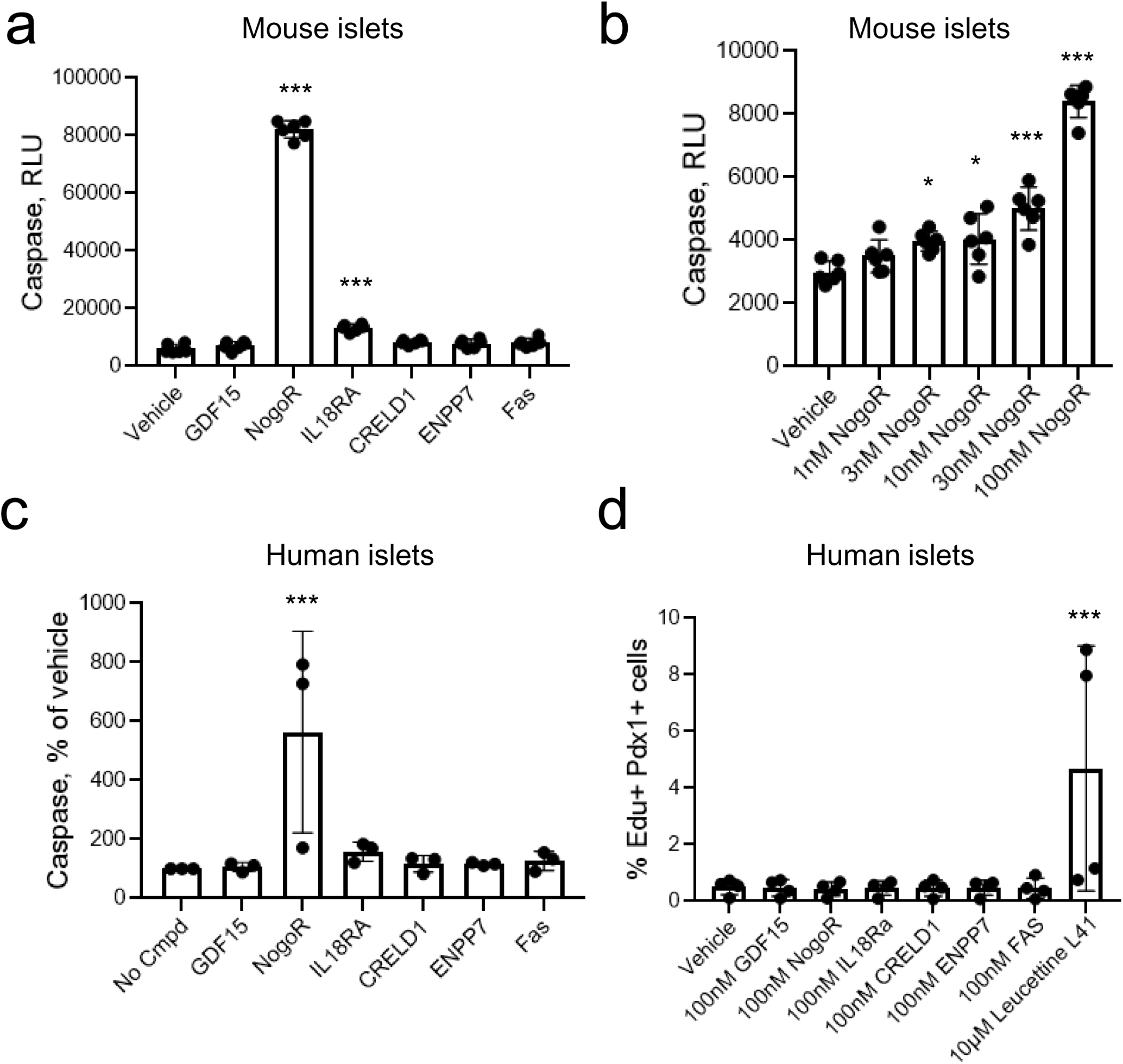
Impact of identified biomarkers on apoptosis (a-c) or proliferation (d) in mouse or human islets as indicated. Test compounds were added at 100 nM unless otherwise indicated. *,***p<0.05, 0.001 by one-way ANOVA for the effects of the indicated compounded *versus* vehicle. See Methods for other details.

### NogoR improves glucose tolerance in vivo

GDF15/MIC has been the subject of several earlier studies (see Discussion) and so was not pursued further here. Since NogoR was the biomarker with the next-largest correlation with disease progression, we next sought to determine whether this protein may influence glucose homeostasis in vivo. Administered daily for two weeks to mice previously maintained on a HFHS diet (four weeks^28^), we observed a clear improvement in glucose tolerance in vivo versus vehicle-injected animals (**Fig. 6a-d**). These changes were observed with no change in insulin secretion (**Fig. 6e**).

**Figure 6.**
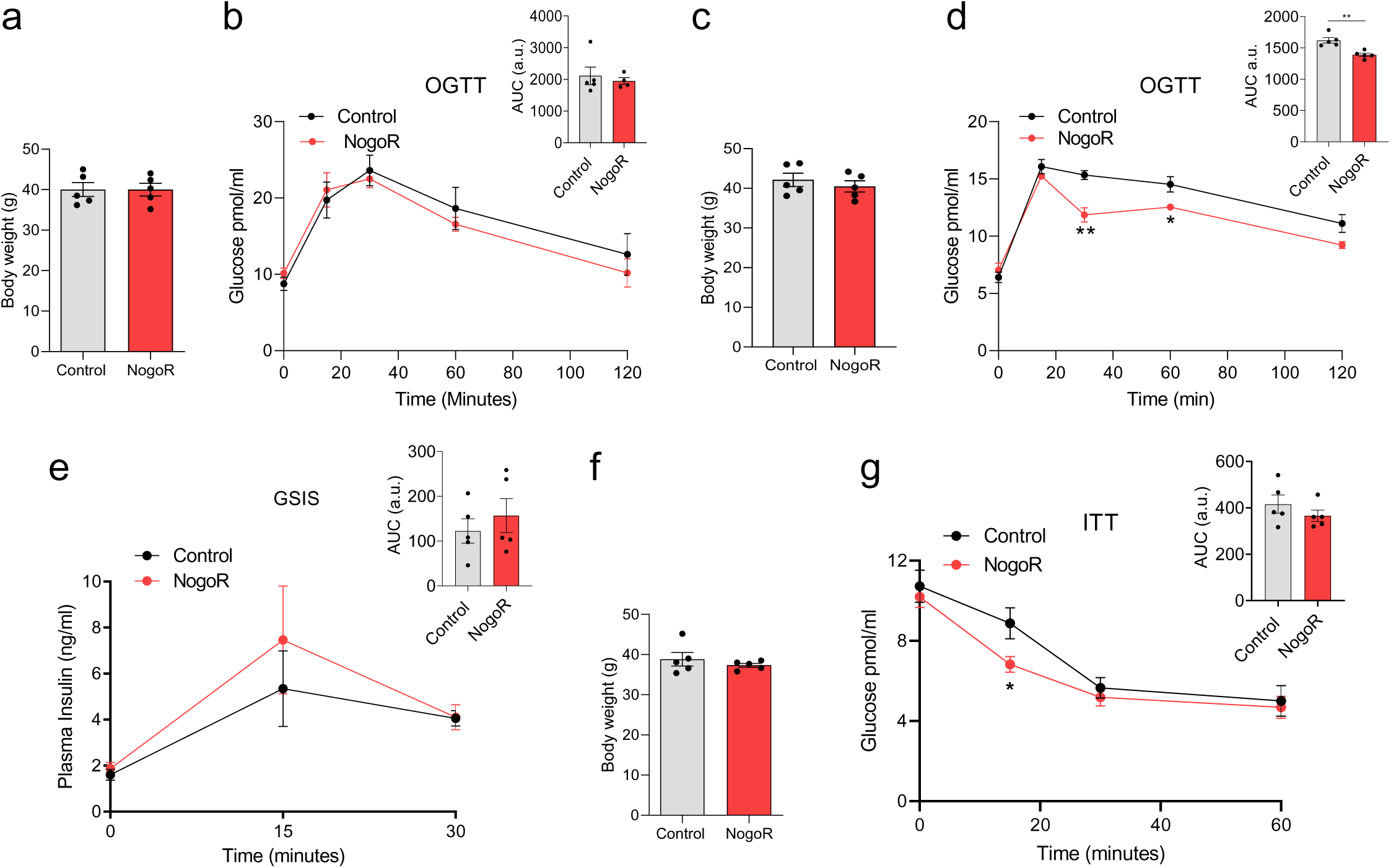
**NogoR enhances glucose clearance and insulin sensitivity in mice**. Two separate cohorts of wild-type male C57BL/6J mice were maintained on a high fat diet for six weeks, then injected for 14 consecutive days with saline or 100 ng (2.1 pmol/animal) recombinant NogoR. **a,b)** Body weights of cohort one and circulating glucose levels during an oral glucose tolerance test (OGTT;2g/kg) pre-NogoR treatment *n* =5. **c,d)** Body weights and blood glucose levels after an oral glucose load (2g/kg) of cohort one after NogoR treatment. *n* = 5. **e)** Plasma insulin levels after an oral glucose load (2g/kg) in cohort 1. *n* = 5 per group. **f, g)** Post-treatment body weights of cohort 2 and circulating glucose levels after receiving an intraperitoneal injection of 1 IU/kg of insulin. Data are represented as the mean ± SEM. *p-value < 0.05 Student T-test.

Since these results suggested that NogoR might affect insulin signalling in disease relevant tissues such as the liver, we measured the action of this protein on signalling events downstream to insulin receptor activation, tested in HepG2 cells. Although not achieving statistical significance, a trend was observed towards a potentiation of insulin-stimulated phosphorylation of the protein kinase AKT on Ser473 after culture at different concentrations of NogoR (1 nM, 10 nM and 100 nM) for 3h. (**SFig. 1a**). In contrast, no differences were observed in insulin-stimulated Akt phosphorylation in HepG2 cells when cultured with different concentrations of CRELD1 (**SFig. 1b**).

### Effect of IL-18Ra on IL-18Ra signalling

After NogoR, IL-18Ra exerted the third strongest impact on diabetes progression (**Fig. 3**). We therefore tested the effects of IL-18Ra in a reporter cell line expressing the IL-18R and a luciferase construct under the control of the cytokine-regulated transcription factor, nuclear factor κB (NF-κB; Methods). IL-18Ra attenuated the actions of IL-18 over a range of concentrations at concentrations as low as 0.1 nM (**Fig. 7**).

**Figure 7.**
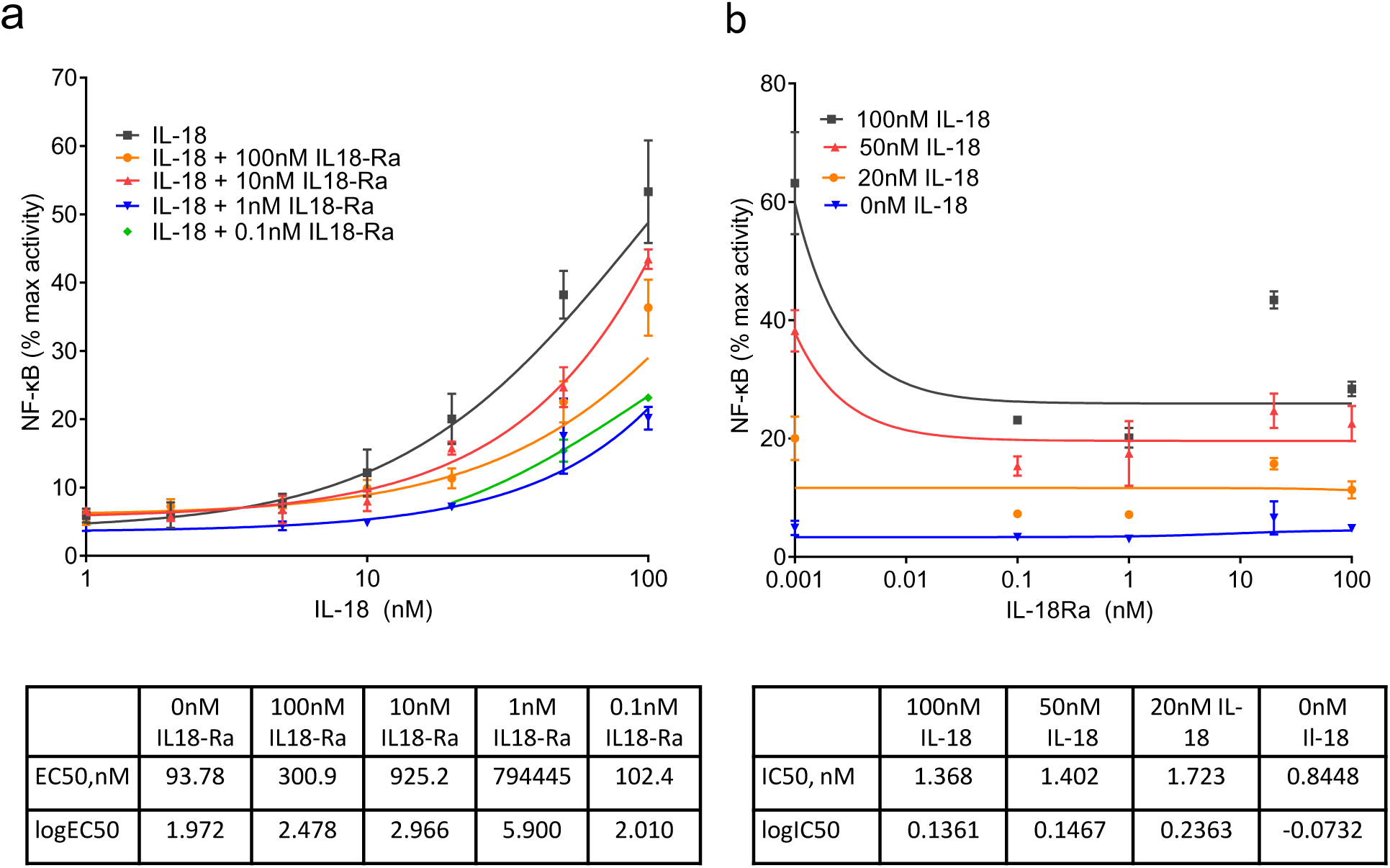
**NF-κB activation in HEK293 cells overexpressing IL-18R and co-stimulated with Il-18 and IL-18Rα.** IL-18R HEK cells stimulated for 6 h with 100, 50, 20, 10, 5, 2 and 1 nM of IL-18 alone, and also with 100, 10, 1 and 0.1nM of IL-18Rα. IL-18 and IL-18Rα concentrations were tested in triplicate and NF-κB activation was measured using dual luciferase reporter assay. Curve fitting was done using non-linear regression with GraphPad Prism 9.0.0 for log(agonist) vs. response (a) and inhibitor vs. response (b).

## DISCUSSION

We have undertaken a large multi-omic study, across three patient cohorts to discover novel lipid, metabolite and protein biomarkers for diabetes progression. Many of our findings are replicated in independent diabetes progression cohorts or validated for incident and/or prevalent diabetes. In particular, we identify 9 lipids, 3 small charged molecules and 11 protein biomarkers associated with accelerated glycaemic deterioration, and provide biological data in pre-clinical models demonstrating possible mechanisms of action for NogoR and IL-18Ra. Strikingly, measurements of proteins and lipid data reveal that the drivers of diabetes incidence and prevalence may be similar to those of progression.

### Metabolites and glycaemic deterioration

Two metabolites, Aminoadipic Acid (AADA) and Homocitrulline (Hcit) were significantly associated with diabetes progression, after correcting for multiple testing; three metabolites, Isoleucine (Ile), and the bile acids GCA and TCA, were nominally associated with progression. Isoleucine is a branched chain amino acid (BCAA) and a well-established risk factor for insulin resistance and increased risk of type 2 diabetes.^29^ Correspondingly, Ile was also associated with both prevalent and incident diabetes in the current study. GCA and TCA are both existing markers for pre-existing diabetes.^30^

In previous studies, AADA was shown to be a risk factor for incident diabetes^27^ and Hcit with prevalent diabetes^31^, consistent with our findings. AADA is an alpha amino acid formed as a downstream product of lysine oxidation by the action of myeloperoxidase (MPO).^32^ Higher levels of plasma AADA have been associated with obesity, insulin resistance, and increased risk of diabetes.^27, 33–35^ Hcit levels have been associated with disrupted energy metabolism in rat brains and have been linked to chronic renal failure.^36, 37^ Of note, however, where there was a sufficient genetic instrument, none of the metabolites we assessed with mendelian randomisation were causally associated with diabetes risk, although it should be noted that BCAAs have been suggested to be causal in the aetiology of type 2 diabetes.^38^

### Lipids and glycaemic deterioration

Nine lipids were associated with diabetes progression of which eight were associated with increased risk which all belonged to the Triacylglycerol (TAG) class. In the external validation data, all eight TAGs were associated with increased risk of type 2 diabetes. TAGs have previously been associated with incident T2D risk.^39^ TAG species levels also strongly decline when obese individuals with diabetes undergo Roux-en-Y gastric bypass.^40^ SM 42:2;2 was the sole lipid associated with lower risk on progression towards to insulin, but was associated with increased risk on future diabetes in the external validation data. A possible explanation for this could be that metformin treatment influences the sphingomyelin levels including SM 42:2;2 as has been shown in two studies in metformin treated HFD animals and human hepatocytes respectively.^41, 42^ The top associated lipids were also investigated for causality but generally instruments were not available or no evidence was observed. The MR analysis of PE 18:0;0_18:2;0 supported a possible causal relation with incident diabetes.

### Proteins and glycaemic deterioration

The protein with the strongest association with time to insulin was GDF-15/MIC1. This protein has previously been implicated in diabetes incidence and the control of food intake^43, 44^, acting via receptors in the hind brain.^28^ GDF-15 has also been reported to serve as a useful biomarker for impaired fasting glucose^45^ and diabetic kidney disease^46^ as well as a number of conditions including cardiovascular disease^47, 48^ GDF15 is strongly elevated following metformin exposure, due to release from the gut.^49^ Of note, we did not show any attenuation of the GDF15 signal with progression when adjusting for whether the patients were treated with metformin at the time of blood sampling.

In addition, we identify several novel proteins biomarkers to be associated with glycaemic deterioration in diabetes, including NogoR (RTN4), IL-18Ra, CRELD1, ENPP7 and FAS. Interestingly, NogoR has previously been associated with prevalent diabetes and diabetes incidence.^13^ In an effort to understand the potential impact of an increase in circulating NogoR on glucose metabolism, we demonstrated that injection of this biomarker improves glucose tolerance in high fat-high sucrose-fed mice, an effect likely to reflect improved insulin sensitivity; insulin secretion was not significantly affected (indeed tended to be *increased* after treatment). NogoR, (encoded by the *RTN4R* gene), is chiefly expressed in the central nervous exist and mediates interactions between the myelin sheath and neurons, and has been implicated in Alzheimer’s disease.^50^ In this setting, NogoR interacts with Oligodendrocyte myelin glycoprotein (OMGP) and Nogo-A, present on myelin cells, to inhibit neuronal regeneration.^51^ Thus NogoR serves as a receptor for Nogo-A but conceivably also the shorter homologues Nogo-B and Nogo-C.^52^ Importantly in the context of systemic glucose homeostasis, Nogo-B interacts with the Nogo-B receptor (NGBR, encoded by *NUS1*) and knockout of *Nus1* in mice causes hepatic steatosis, possibly be interfering with insulin signalling.^53^ Furthermore, variants in the human *NUS1* gene including rs4443534 are associated with altered type 2 diabetes risk.^54^ Thus, by titrating Nogo-B (or other Nogo family members) circulating NogoR may act indirectly on the liver to affect glucose storage or glycogen breakdown. On the other hand, NogoR also enhanced cell death in pancreatic islets, at least at high concentrations of this biomarker. The mechanisms involved in this action are unclear given the absence of expression of the parental receptors of the interactors Nogo-A and OMGP in islets.^55^ Furthermore, the physiological relevance of these changes is uncertain since concentrations at which NogoR exerted these effects in vitro (≥ 3 nM) comfortably exceeded those which improved glucose metabolism in vivo (∼ 0.7 nM). We suspect, therefore, that positive effects of NogoR to improve insulin sensitivity may predominant in most physiological settings, though we do not exclude a shift towards deleterious actions on the pancreas in disease settings.

CRELD1 (Cysteine-Rich with EGF-Like Domains 1, AVSD2, Cirrin) is a membrane-bound Ca^2+^-binding member of the EGF family critically required for normal development of the heart^56^ whose expression was recently shown to influence T-cell activity and immune homeostasis.^57^ Although we were not able to test the effects of this agent on glucose homeostasis in vivo due to the unavailability of the human protein, we do provide evidence that CRELD1 may regulate insulin signalling *in vitro*.

A recent Mendelian randomisation study^13^ provided nominally significant support for a causal association between IL-18Ra and T2D, which was repeated here. Consistent with a role in glucose homeostasis, IL-18 deletion in the mouse leads to obesity and insulin resistance.^58^ Conversely, after weight loss following exercise/diet or bariatric surgery in man, a significant reduction in IL-18 concentrations was observed.^59, 60^ IL-18 secretion also increased in response to inflammasome activation and pyroptosis.^61^ We confirmed earlier studies^62^ which demonstrated that an IL-18Ra:Fc fusion fragment inhibits the pro-inflammatory action of IL-18. However, and in distinction to these earlier studies, measuring interferon-gamma (IFN-gamma) production from mononuclear cells, the actions of IL-18Ra did not depend upon the additional presence of IL-18Rbeta. We also noted that the concentrations of IL-18Ra as low as 0.1 nM efficiently inhibited the actions of a considerable excess (100 nM) of IL-18 potentially suggesting a non-competitive action (*i.e.*, binding to the IL-18R at a separate site on the cellular receptor to IL-18). Unexpectedly, IL-18Ra also influenced beta cell apoptosis in vitro. The mechanisms involved here are unclear, however, since IL-18Ra expression levels in the beta cell are low.^63^ Finally, we note that IL-18Ra, CRELD1 and coactosin-like protein are all involved in immune regulation, and might thus influence the inflammatory changes known to be involved in T2D.^64^

ENPP7 (Ectonucleotide pyrophosphatase/phosphodiesterase-7) is strongly expressed in the small intestine where it is involved in sphingomyelin hydrolysis and the absorption of ceramide and phosphocholine.^65^ These processes might therefore be influenced by changed circulating levels of ENPP7. FAS (FAS cell surface death receptor; TNF superfamily receptor-6) is involved in caspase 3 and 8 activation and cell death^66^ and might thus influence cell survival in critical metabolic tissues. HSP 90B (heat shock protein 90 alpha family class B member 1), encoded by *HSP90AB1,* is a molecular chaperone and regulator of protein folding.^67^

Other identified proteins were associated with protection from glycaemic deterioration of diabetes. SMAC/IAPP-binding mitochondrial protein, also called DIABLO, is a mitochondrially-associated protein which migrates to the cytosol upon the activation of apoptosis, facilitating this process by restricting the activity of apoptotic inhibitors.^68^ Lowered levels of this protein in the plasma thus seem likely to reflect diminished levels of cell death in disease-relevant (or other) tissues. Coactosin-like protein, encoded by the *COTL1* gene and also called Coactosin-like F-actin binding protein, and CLP, is enriched in haematopoietic cells (BioGPS), and regulates leukotriene synthesis. Low levels may therefore reflect more limited inflammation in individuals whose disease deteriorates quickly. Testican-1, also called SPOCK1, SPARC, and Osteonectin is enriched in the brain. The function of SPOCK1, a Ca^2+^ binding proteoglycan, is currently unknown, though roles in neuronal development^69^ adipocyte differentiation^70^ and as an extracellular matrix factor controlling epithelial to mesenchymal transition^71^ have been suggested. Finally, HEMK2 (N6AMT1, PrmC) is a ubiquitously-expressed DNA methyl transferase.^72^

To investigate causal associations, we undertook MR analysis of six of our identified protein biomarkers that were associated with diabetes progression. We repeated previous findings that GDF15 is causally associated with diabetes progression^13^ and found a possible causal association for IL-18Ra and FAS. These data suggest that these two proteins are likely to be causally associated with diabetes progression. The lack of a causal association of NogoR with diabetes risk does suggest that NogoR may similarly not be causally associated with diabetes progression, however, our functional studies do suggest a causal mechanism linking NogoR with abnormal glucose metabolism.

This study has several limitations. Between cohorts there was heterogeneity, and this was in part due to the nature of the cohorts, for example the ACCELERATE cohort is a clinical trial which is different in setup than the discovery studies. Nonetheless, the results of the study were robust, for example a large number of biomarkers were also found to be associated with prevalent and incident diabetes. A third limitation is that for some protein biomarkers ELISA assays are currently unavailable to validate the signals obtained in the SomaLogic screen. Finally, in the Mendelian Randomization analysis we could only investigate the causal association with diabetes risk, and some of the genetic instruments were weak, so the absence of causality in MR analysis does not mean that our findings with diabetes progression were non-causal. Indeed, plausible evidence for causality was subsequently obtained in functional studies in preclinical models for NogoR and IL-18Ra.

## CONCLUSION

Our findings provide new mechanistic insights into the mechanisms that underlie glycaemic deterioration once diabetes has developed. Importantly, we describe novel biomarkers of different chemical classes, several of which have not previously been associated with the incident or prevalent disease, suggestive of potentially distinct mechanisms driving the two stages of glycaemic deterioration - towards diabetes onset and after diabetes onset. We provide direct functional analyses implicating two of these (NogoR, IL-18Ra) as likely to contribute directly to disease progression. By better understanding the biological drivers of glycaemic deterioration in diabetes it may be possible to target therapies to these processes to prevent or slow diabetes progression, rather than simply to lower HbA1c, potentially transforming diabetes treatments.

## Data Availability

Summary statistics of lipidomic, proteomic and metabolomic data will be available from an interactive Shiny dashboard available upon publication.

## ACKNOWLEDGEMENTS

We acknowledge the support of the Health Informatics Centre, University of Dundee for managing and supplying the anonymised data. The authors wish to thank the Core-IT group of SIB Swiss Institute of Bioinformatics and in particular Jorge Molina for expert technical help in setting up and maintaining the federated database. GR thanks Claudio Elgueta Karstegl for technical support. The authors thank participants of the included cohorts.

## AUTHOR CONTRIBUTION

RCS, LAD, JWJB, LMTH, ERP, and GR designed the study. GR and RCS drafted the manuscript. RCS, LAD, HF, GAB, MA performed the analyses. GR oversaw biomarker shipments and assays, and coordinated all functional work in preclinical models. MS, MB, LMtH, AAWAvH, PJME, JWJB, EA, LG, LAD, ERP provided patient samples for analysis. LL, HM, EG, IL performed all studies on NogoR, EA, MS work on IL-18Ra. ID, DK, FB, DM, AN, MI set up a federated node system for data-analysis. RCS, LAD, HF, DMA, EA, AA, MJG, MK, FM, TS, AW, CLQ, MI were involved in the (clinical) data pre-processing and quality control. GNG, AF, MKH, DMA, IP, TJP, BT, VL, LG, PWF, GAR contributed to the data acquisition and project logistics. MJG, CK, KS generated the Lipotype data. CLQ, AA, PR, AW, TS, FM generated the metabolomics data and/or were involved in the quality control. FO, CF and OM acquired the MDC-CC data and performed the lipid and protein validation in the MDC-CC. MC and PF performed the metabolite validation in DESIR. VaG, ViG and LLJ performed the protein validation in the AGES-Reykjavik cohort. PJME and AAWAvH acquired the data from the Hoorn DCS cohort. All authors contributed to the data interpretation. All authors critically revised the manuscript and approved the final version. RCS and LAD are the guarantors of the work.

## SUPPLEMENTARY FIGURE LEGENDS

**Supp Figure 1.** Effects of NogoR and of CRELD1 on insulin signalling in vitro. HepG2 cells were cultured for 3 hours with 0, 1, 10 or 100 nM of NogoR (a) or CRELD1 (b) and 100 nM of insulin for 15 minutes prior protein extraction. a) Protein AKT and pSer473 AKT levels in Hepg2 treated with different concentrations of NogoR (*n* = 3). b) Protein AKT and pSer473 AKT levels in HepG2 treated with different concentrations of NogoR (*n* = 3). Data are represented as the mean ± SEM and were analysed by one-way ANOVA.

