## Supplementary figures and images for "Novel biomarkers for glycaemic deterioration in type 2 diabetes: an IMI RHAPSODY study"

### Supp. Fig. 1

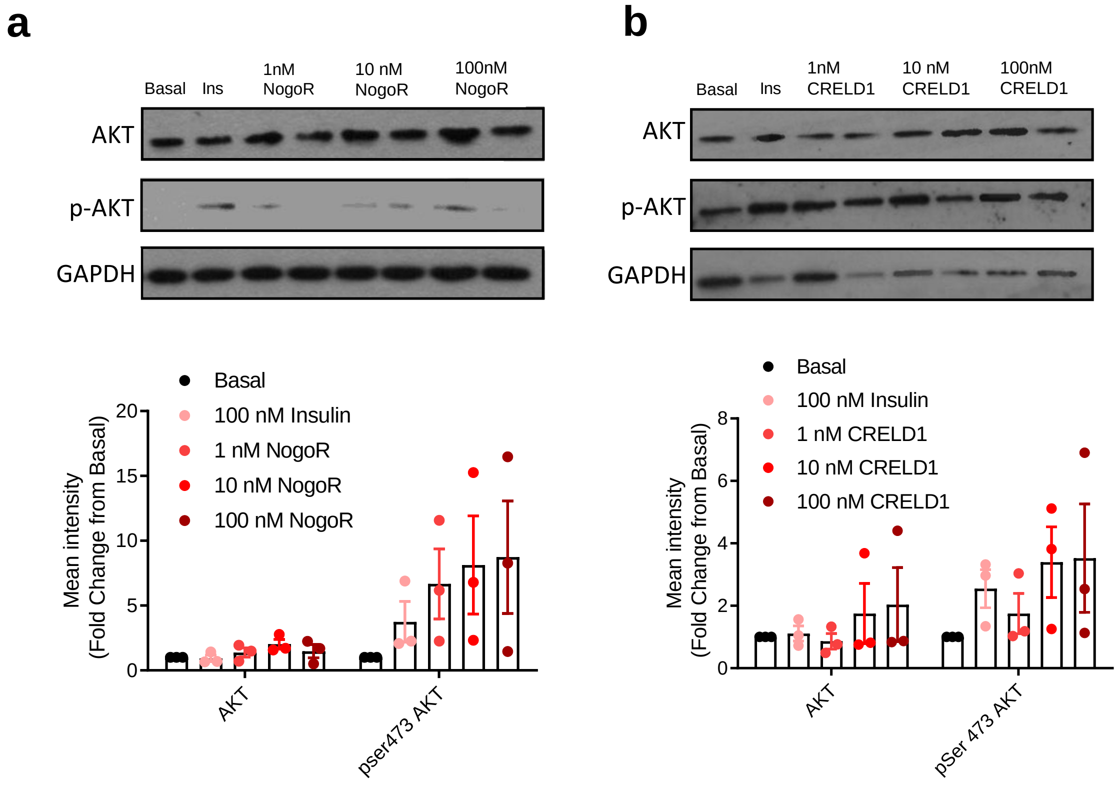
