## Supplementary material for "Novel biomarkers for glycaemic deterioration in type 2 diabetes: an IMI RHAPSODY study": Supp Tables S1-S3

**Table S1 Included lipid species with SwissLipids names**

| **feature** | **SwissLipids Name** | **SwissLipids ID** |
| --- | --- | --- |
| CE 14:0;0 | Sterol ester (27:1/14:0) | SLM:000500342 |
| CE 15:0;0 | Sterol ester (27:1/15:0) | SLM:000500343 |
| CE 16:0;0 | Sterol ester (27:1/16:0) | SLM:000500346 |
| CE 16:1;0 | Sterol ester (27:1/16:1) | SLM:000500345 |
| CE 17:0;0 | Sterol ester (27:1/17:0) | SLM:000500347 |
| CE 17:1;0 | Sterol ester (27:1/17:1) |  |
| CE 18:0;0 | Sterol ester (27:1/18:0) | SLM:000500352 |
| CE 18:1;0 | Sterol ester (27:1/18:1) | SLM:000500351 |
| CE 18:2;0 | Sterol ester (27:1/18:2) | SLM:000500350 |
| CE 18:3;0 | Sterol ester (27:1/18:3) | SLM:000500349 |
| CE 20:2;0 | Sterol ester (27:1/20:2) | SLM:000500357 |
| CE 20:3;0 | Sterol ester (27:1/20:3) | SLM:000500356 |
| CE 20:4;0 | Sterol ester (27:1/20:4) | SLM:000500355 |
| CE 20:5;0 | Sterol ester (27:1/20:5) | SLM:000500354 |
| CE 22:6;0 | Sterol ester (27:1/22:6) | SLM:000500361 |
| Cer 40:1;2 | Ceramide (d40:1) | SLM:000391319 |
| Cer 40:2;2 | Ceramide (d40:2) | SLM:000391317 |
| Cer 42:1;2 | Ceramide (d42:1) | SLM:000391346 |
| Cer 42:2;2 | Ceramide (d42:2) | SLM:000391345 |
| Chol | cholesterol | SLM:000000287 |
| DAG 16:0;0_18:1;0 | Diacylglycerol (16:0_18:1) | SLM:000308862 |
| DAG 16:0;0_18:2;0 | Diacylglycerol (16:0_18:2) | SLM:000308863 |
| DAG 16:1;0_18:1;0 | Diacylglycerol (16:1_18:1) | SLM:000308894 |
| DAG 18:1;0_18:1;0 | Diacylglycerol (18:1_18:1) | SLM:000309012 |
| DAG 18:1;0_18:2;0 | Diacylglycerol (18:1_18:2) | SLM:000309013 |
| DAG 18:1;0_18:3;0 | Diacylglycerol (18:1_18:3) | SLM:000309014 |
| DAG 18:2;0_18:2;0 | Diacylglycerol (18:2_18:2) | SLM:000309040 |
| LPC 16:0;0 | Phosphatidylcholine (16:0_0:0) | SLM:000063723 |
| LPC 16:1;0 | Phosphatidylcholine (16:1_0:0) | SLM:000063777 |
| LPC 18:0;0 | Phosphatidylcholine (18:0_0:0) | SLM:000063933 |
| LPC 18:1;0 | Phosphatidylcholine (18:1_0:0) | SLM:000063983 |
| LPC 18:2;0 | Phosphatidylcholine (18:2_0:0) | SLM:000064032 |
| LPC 20:3;0 | Phosphatidylcholine (20:3_0:0) | SLM:000064347 |
| LPC 20:4;0 | Phosphatidylcholine (20:4_0:0) | SLM:000064388 |
| LPE 18:1;0 | Phosphatidylethanolamine (18:1_0:0) | SLM:000067947 |
| LPE 18:2;0 | Phosphatidylethanolamine (18:2_0:0) | SLM:000067996 |
| LPE 20:4;0 | Phosphatidylethanolamine (20:4_0:0) | SLM:000068352 |
| PC 14:0;0_18:1;0 | Phosphatidylcholine (14:0_18:1) | SLM:000063564 |
| PC 14:0;0_18:2;0 | Phosphatidylcholine (14:0_18:2) | SLM:000063565 |
| PC 16:0;0_16:0;0 | Phosphatidylcholine (16:0_16:0) | SLM:000063724 |
| PC 16:0;0_16:1;0 | Phosphatidylcholine (16:0_16:1) | SLM:000063725 |
| PC 16:0;0_17:1;0 | Phosphatidylcholine (16:0_17:1) |  |
| PC 16:0;0_18:0;0 | Phosphatidylcholine (16:0_18:0) | SLM:000063728 |
| PC 16:0;0_18:1;0 | Phosphatidylcholine (16:0_18:1) | SLM:000063729 |
| PC 16:0;0_18:2;0 | Phosphatidylcholine (16:0_18:2) | SLM:000063730 |
| PC 16:0;0_18:3;0 | Phosphatidylcholine (16:0_18:3) | SLM:000063731 |
| PC 16:0;0_20:1;0 | Phosphatidylcholine (16:0_20:1) | SLM:000063735 |
| PC 16:0;0_20:2;0 | Phosphatidylcholine (16:0_20:2) | SLM:000063736 |
| PC 16:0;0_20:3;0 | Phosphatidylcholine (16:0_20:3) | SLM:000063737 |
| PC 16:0;0_20:4;0 | Phosphatidylcholine (16:0_20:4) | SLM:000063738 |
| PC 16:0;0_20:5;0 | Phosphatidylcholine (16:0_20:5) | SLM:000063739 |
| PC 16:0;0_22:4;0 | Phosphatidylcholine (16:0_22:4) | SLM:000063745 |
| PC 16:0;0_22:5;0 | Phosphatidylcholine (16:0_22:5) | SLM:000063746 |
| PC 16:0;0_22:6;0 | Phosphatidylcholine (16:0_22:6) | SLM:000063747 |
| PC 16:1;0_18:1;0 | Phosphatidylcholine (16:1_18:1) | SLM:000063782 |
| PC 16:1;0_18:2;0 | Phosphatidylcholine (16:1_18:2) | SLM:000063783 |
| PC 17:0;0_18:1;0 | Phosphatidylcholine (17:0_18:1) | SLM:000063885 |
| PC 17:0;0_18:2;0 | Phosphatidylcholine (17:0_18:2) | SLM:000063886 |
| PC 17:0;0_20:3;0 | Phosphatidylcholine (17:0_20:3) | SLM:000063893 |
| PC 17:0;0_20:4;0 | Phosphatidylcholine (17:0_20:4) | SLM:000063894 |
| PC 18:0;0_18:1;0 | Phosphatidylcholine (18:0_18:1) | SLM:000063935 |
| PC 18:0;0_18:2;0 | Phosphatidylcholine (18:0_18:2) | SLM:000063936 |
| PC 18:0;0_18:3;0 | Phosphatidylcholine (18:0_18:3) | SLM:000063937 |
| PC 18:0;0_20:3;0 | Phosphatidylcholine (18:0_20:3) | SLM:000063943 |
| PC 18:0;0_20:4;0 | Phosphatidylcholine (18:0_20:4) | SLM:000063944 |
| PC 18:0;0_20:5;0 | Phosphatidylcholine (18:0_20:5) | SLM:000063945 |
| PC 18:0;0_22:5;0 | Phosphatidylcholine (18:0_22:5) | SLM:000063952 |
| PC 18:0;0_22:6;0 | Phosphatidylcholine (18:0_22:6) | SLM:000063953 |
| PC 18:1;0_18:1;0 | Phosphatidylcholine (18:1_18:1) | SLM:000063984 |
| PC 18:1;0_18:2;0 | Phosphatidylcholine (18:1_18:2) | SLM:000063985 |
| PC 18:1;0_20:3;0 | Phosphatidylcholine (18:1_20:3) | SLM:000063992 |
| PC 18:1;0_20:4;0 | Phosphatidylcholine (18:1_20:4) | SLM:000063993 |
| PC 18:2;0_18:2;0 | Phosphatidylcholine (18:2_18:2) | SLM:000064033 |
| PC 18:2;0_20:4;0 | Phosphatidylcholine (18:2_20:4) | SLM:000064041 |
| PC O-16:0;0/16:0;0 | Phosphatidylcholine (O-16:0_16:0) | SLM:000065919 |
| PC O-16:0;0/16:1;0 | Phosphatidylcholine (O-16:0_16:1) | SLM:000065920 |
| PC O-16:0;0/18:1;0 | Phosphatidylcholine (O-16:0_18:1) | SLM:000065924 |
| PC O-16:0;0/18:2;0 | Phosphatidylcholine (O-16:0_18:2) | SLM:000065925 |
| PC O-16:0;0/20:3;0 | Phosphatidylcholine (O-16:0_20:3) | SLM:000065932 |
| PC O-16:0;0/20:4;0 | Phosphatidylcholine (O-16:0_20:4) | SLM:000065933 |
| PC O-16:1;0/16:0;0 | Phosphatidylcholine (O-16:1_16:0) | SLM:000065984 |
| PC O-16:1;0/18:0;0 | Phosphatidylcholine (O-16:1_18:0) | SLM:000065988 |
| PC O-16:1;0/18:1;0 | Phosphatidylcholine (O-16:1_18:1) | SLM:000065989 |
| PC O-16:1;0/18:2;0 | Phosphatidylcholine (O-16:1_18:2) | SLM:000065990 |
| PC O-16:1;0/20:4;0 | Phosphatidylcholine (O-16:1_20:4) | SLM:000065998 |
| PC O-17:0;0/17:1;0 | Phosphatidylcholine (O-17:0_17:1) |  |
| PC O-18:0;0/14:0;0 | Phosphatidylcholine (O-18:0_14:0) | SLM:000066176 |
| PC O-18:0;0/20:4;0 | Phosphatidylcholine (O-18:0_20:4) | SLM:000066193 |
| PC O-18:1;0/16:0;0 | Phosphatidylcholine (O-18:1_16:0) | SLM:000066244 |
| PC O-18:1;0/18:2;0 | Phosphatidylcholine (O-18:1_18:2) | SLM:000066250 |
| PC O-18:1;0/20:3;0 | Phosphatidylcholine (O-18:1_20:3) | SLM:000066257 |
| PC O-18:1;0/20:4;0 | Phosphatidylcholine (O-18:1_20:4) | SLM:000066258 |
| PC O-18:2;0/16:0;0 | Phosphatidylcholine (O-18:2_16:0) | SLM:000066309 |
| PC O-18:2;0/18:1;0 | Phosphatidylcholine (O-18:2_18:1) | SLM:000066314 |
| PC O-18:2;0/18:2;0 | Phosphatidylcholine (O-18:2_18:2) | SLM:000066315 |
| PE 16:0;0_18:2;0 | Phosphatidylethanolamine (16:0_18:2) | SLM:000067694 |
| PE 16:0;0_20:4;0 | Phosphatidylethanolamine (16:0_20:4) | SLM:000067702 |
| PE 18:0;0_18:2;0 | Phosphatidylethanolamine (18:0_18:2) | SLM:000067900 |
| PE 18:0;0_20:4;0 | Phosphatidylethanolamine (18:0_20:4) | SLM:000067908 |
| PE 18:1;0_18:1;0 | Phosphatidylethanolamine (18:1_18:1) | SLM:000067948 |
| PE O-16:1;0/20:4;0 | Phosphatidylethanolamine (O-16:1_20:4) | SLM:000069962 |
| PE O-18:1;0/18:2;0 | Phosphatidylethanolamine (O-18:1_18:2) | SLM:000070214 |
| PE O-18:1;0/20:4;0 | Phosphatidylethanolamine (O-18:1_20:4) | SLM:000070222 |
| PE O-18:2;0/18:1;0 | Phosphatidylethanolamine (O-18:2_18:1) | SLM:000070278 |
| PE O-18:2;0/18:2;0 | Phosphatidylethanolamine (O-18:2_18:2) | SLM:000070279 |
| PE O-18:2;0/20:4;0 | Phosphatidylethanolamine (O-18:2_20:4) | SLM:000070287 |
| PI 16:0;0_18:1;0 | Phosphatidylinositol (16:0_18:1) | SLM:000073801 |
| PI 16:0;0_18:2;0 | Phosphatidylinositol (16:0_18:2) | SLM:000073802 |
| PI 18:0;0_18:1;0 | Phosphatidylinositol (18:0_18:1) | SLM:000074007 |
| PI 18:0;0_18:2;0 | Phosphatidylinositol (18:0_18:2) | SLM:000074008 |
| PI 18:0;0_20:3;0 | Phosphatidylinositol (18:0_20:3) | SLM:000074015 |
| PI 18:0;0_20:4;0 | Phosphatidylinositol (18:0_20:4) | SLM:000074016 |
| PI 18:1;0_18:1;0 | Phosphatidylinositol (18:1_18:1) | SLM:000074056 |
| PI 18:1;0_18:2;0 | Phosphatidylinositol (18:1_18:2) | SLM:000074057 |
| SM 32:1;2 | Sphingomyelin (d32:1) | SLM:000390695 |
| SM 34:0;2 | Sphingomyelin (d34:0) | SLM:000390716 |
| SM 34:1;2 | Sphingomyelin (d34:1) | SLM:000390714 |
| SM 34:2;2 | Sphingomyelin (d34:2) | SLM:000390712 |
| SM 36:1;2 | Sphingomyelin (d36:1) | SLM:000390739 |
| SM 36:2;2 | Sphingomyelin (d36:2) | SLM:000390737 |
| SM 38:2;2 | Sphingomyelin (d38:2) | SLM:000390765 |
| SM 40:1;2 | Sphingomyelin (d40:1) | SLM:000390797 |
| SM 40:2;2 | Sphingomyelin (d40:2) | SLM:000390795 |
| SM 42:2;2 | Sphingomyelin (d42:2) | SLM:000390823 |
| TAG 46:1;0 | Triacylglycerol (46:1) | SLM:000308244 |
| TAG 46:2;0 | Triacylglycerol (46:2) | SLM:000308245 |
| TAG 48:0;0 | Triacylglycerol (48:0) | SLM:000308257 |
| TAG 48:1;0 | Triacylglycerol (48:1) | SLM:000308258 |
| TAG 48:2;0 | Triacylglycerol (48:2) | SLM:000308259 |
| TAG 48:3;0 | Triacylglycerol (48:3) | SLM:000308260 |
| TAG 49:1;0 | Triacylglycerol (49:1) | SLM:000308267 |
| TAG 49:2;0 | Triacylglycerol (49:2) | SLM:000308268 |
| TAG 50:1;0 | Triacylglycerol (50:1) | SLM:000308276 |
| TAG 50:2;0 | Triacylglycerol (50:2) | SLM:000308277 |
| TAG 50:3;0 | Triacylglycerol (50:3) | SLM:000308278 |
| TAG 50:4;0 | Triacylglycerol (50:4) | SLM:000308279 |
| TAG 50:5;0 | Triacylglycerol (50:5) | SLM:000308280 |
| TAG 51:1;0 | Triacylglycerol (51:1) | SLM:000308286 |
| TAG 51:2;0 | Triacylglycerol (51:2) | SLM:000308287 |
| TAG 51:3;0 | Triacylglycerol (51:3) | SLM:000308288 |
| TAG 51:4;0 | Triacylglycerol (51:4) | SLM:000308289 |
| TAG 52:2;0 | Triacylglycerol (52:2) | SLM:000308298 |
| TAG 52:3;0 | Triacylglycerol (52:3) | SLM:000308299 |
| TAG 52:4;0 | Triacylglycerol (52:4) | SLM:000308300 |
| TAG 52:5;0 | Triacylglycerol (52:5) | SLM:000308301 |
| TAG 52:6;0 | Triacylglycerol (52:6) | SLM:000308302 |
| TAG 53:2;0 | Triacylglycerol (53:2) | SLM:000308309 |
| TAG 53:3;0 | Triacylglycerol (53:3) | SLM:000308310 |
| TAG 53:4;0 | Triacylglycerol (53:4) | SLM:000308311 |
| TAG 54:3;0 | Triacylglycerol (54:3) | SLM:000308323 |
| TAG 54:4;0 | Triacylglycerol (54:4) | SLM:000308324 |
| TAG 54:5;0 | Triacylglycerol (54:5) | SLM:000308325 |
| TAG 54:6;0 | Triacylglycerol (54:6) | SLM:000308326 |
| TAG 54:7;0 | Triacylglycerol (54:7) | SLM:000308327 |
| TAG 56:3;0 | Triacylglycerol (56:3) | SLM:000308349 |
| TAG 56:4;0 | Triacylglycerol (56:4) | SLM:000308350 |
| TAG 56:5;0 | Triacylglycerol (56:5) | SLM:000308351 |
| TAG 56:6;0 | Triacylglycerol (56:6) | SLM:000308352 |
| TAG 56:7;0 | Triacylglycerol (56:7) | SLM:000308353 |
| TAG 56:8;0 | Triacylglycerol (56:8) | SLM:000308354 |
| TAG 58:7;0 | Triacylglycerol (58:7) | SLM:000308381 |
| TAG 58:8;0 | Triacylglycerol (58:8) | SLM:000308382 |

**Table S2 Characteristics of the included discovery and validation cohorts**

| **Metabolomics** |  |  |  |  |  |
| --- | --- | --- | --- | --- | --- |
|  | *DCS* | *GoDARTS (discovery)* | *GoDARTS (validation)* | *ANDIS (discovery)* | *ANDIS (validation)* |
| N | 1267 | 897 | 699 | 811 | 1969 |
| N events | 227 | 311 | 307 | 74 | 361 |
| %Males | 55.96 | 56.97 | 56.22 | 60.54 | 59.9 |
| Age (years) | 63.83[57.37-70.77] | 62.35[54.5-70.76] | 64.61[56.78-72.67] | 61.89[54.09-69.05] | 62.76[55.53-70.05] |
| BMI (kg/m2) | 30.34[26.7-33.1] | 32.4[28-35.8] | 31.54[27.45-34.6] | 31.71[27.94-34.69] | 30.83[27.08-33.92] |
| HbA1c (mmol/mol) | 47.08[42-50] | 55.54[46-61] | 56.88[48-62] | 60.06[46.0-68.0] | 60.01[44.88-67.75] |
| C-peptide (nmol/L) | 1.15[0.83-1.42] | 2.1[1.35-2.71] | 2.06[1.36-2.52] | 1.32[0.93-1.6] | 1.26[0.86-1.53] |
| HDL (mmol/L) | 1.24[0.99-1.43] | 1.31[1.06-1.5] | 1.32[1.1-1.51] | 1.2[0.96-1.4] | 1.21[0.94-1.4] |
| LDL (mmol/L) | 2.58[1.9-3.2] | 2.15[1.58-2.63] | 2.04[1.45-2.47] | 3.21[2.5-3.9] | 3.06[2.3-3.7] |
| Triglycerides (mmol/L) | 1.79[1.15-2.18] | 2.32[1.4-2.77] | 2.31[1.46-2.76] | 2.09[1.2-2.4] | 2.07[1.2-2.4] |
| Diabetes duration (years) | 2.63[1.43-3.76] | 1.4[0.65-2.16] | 3.95[3.36-4.54] | 0[0-0] | 0[0-0] |
| Metformin (%) | 69.06 | 47.27 | 58.94 | 76.94 | 59.49 |
| Sulfonylureas (%) | 24.07 | 19.06 | 31.18 | 3.08 | 2.89 |
| **Lipidomics** |  |  |  |  |  |
|  | *DCS* | *GoDARTS (discovery)* | *ANDIS (discovery)* |  |  |
| N | 900 | 899 | 809 |  |  |
| N events | 115 | 311 | 71 |  |  |
| %Males | 56.44 | 56.95 | 60.32 |  |  |
| Age (years) | 63.64[57.18-70.34] | 62.32[54.47-70.75] | 61.98[54.44-69.05] |  |  |
| BMI (kg/m2) | 30.22[26.67-33.1] | 32.42[28-35.8] | 31.67[27.93-34.63] |  |  |
| HbA1c (mmol/mol) | 47[41-49.73] | 55.52[46-61] | 60.01[46-68] |  |  |
| C-peptide (nmol/L) | 1.16[0.84-1.42] | 2.1[1.35-2.7] | 1.32[0.93-1.6] |  |  |
| HDL (mmol/L) | 1.24[1-1.44] | 1.31[1.06-1.5] | 1.21[0.96-1.4] |  |  |
| LDL (mmol/L) | 2.61[1.9-3.2] | 2.15[1.58-2.63] | 3.21[2.5-3.9] |  |  |
| Triglycerides (mmol/L) | 1.8[1.14-2.22] | 2.29[1.4-2.75] | 2.03[1.2-2.4] |  |  |
| Diabetes duration (years) | 1.97[1.21-2.78] | 1.4[0.65-2.16] | 0[0-0] |  |  |
| Metformin (%) | 66.78 | 47.27 | 76.88 |  |  |
| Sulfonylureas (%) | 20.56 | 19.13 | 3.09 |  |  |

| **Proteomics** |  |  |  |  |
| --- | --- | --- | --- | --- |
|  | *DCS* | *GoDARTS* | *ANDIS (validation)* | *ACCELERATE (validation)* |
| N | 589 | 599 | 1992 | 1850 |
| N events | 71 | 200 | 222 | 162 |
| %Males | 56.71 | 59.1 | 60.74 | 78.7 |
| Age (years) | 63.19[56.24-70.24] | 61.9[54.09-70.22] | 60.99[54.57-67.96] | 67.17[61.4-72.5] |
| BMI (kg/m2) | 30.25[26.7-33.1] | 32.23[27.7-35.75] | 31.93[28.2-34.9] | 30.0[26.8-33.9] |
| HbA1c (mmol/mol) | 46.84[41-49.73] | 55.44[48-61] | 59.44[45.91-66.71] | 46.0[42.0-54.0] |
| C-peptide (nmol/L) | 1.16[0.84-1.44] | 2.15[1.37-2.76] | 1.36[0.97-1.6] | 3.44[2.52-4.59] |
| HDL (mmol/L) | 1.23[1-1.4] | 1.29[1.06-1.47] | 1.18[0.94-1.4] | 1.14[0.96-1.35] |
| LDL (mmol/L) | 2.65[2-3.3] | 2.19[1.62-2.69] | 3.11[2.4-3.8] | 1.99[1.63-2.43] |
| Triglycerides (mmol/L) | 1.83[1.16-2.24] | 2.32[1.43-2.77] | 2.16[1.2-2.4] | 1.53[1.13-2.06] |
| Diabetes duration (years) | 1.45[1.1-2.06] | 0.92[0.26-1.41] | 0[0-0] | 6.4[3.2-11.4] |
| Metformin (%) | 67.23 | 45.41 | 71.03 | 68.5 |
| Sulfonylureas (%) | 18.68 | 17.20 | 3.16 | 31.6 |

**Table S3 Cox proportional hazard ratio of base models without biomarkers.**

|  | *DCS* |  | *GoDARTS* |  | *ANDIS* |  | *ACCELERATE* |  |
| --- | --- | --- | --- | --- | --- | --- | --- | --- |
| *n* | *3052* |  | *4679* |  | *6068* |  | 1850 |  |
| *n* events | *536* |  | *1256* |  | *723* |  | 161 |  |
| **Model 1 (age, sex, BMI)** | | |  |  |  |  |  |  |
| Variable | *HR* | *P-value* | *HR* | *P-value* | *HR* | *P-value* | *HR* | *P-value* |
| Age | 0.97[0.96-0.98] | 1.64E-15 | 0.96[0.96-0.97] | <0.0001 | 0.99[0.99-1.01] | 0.5648 | 0.98[0.96-0.99] | 0.01 |
| Sex | 1.09[0.91-1.31] | 0.38 | 0.93[0.82-1.04] | 0.1862 | 1.05[0.90-1.23] | 0.5394 | 1.10[0.75-1.61] | 0.62 |
| BMI | 1.00[0.98-1.02] | 0.48 | 1.01[1.00-1.02] | 0.0133 | 0.97[0.96-0.99] | 0.0002 | 1.01[0.99-1.04] | 0.24 |
| **Model 2 (M1 + HDL, C-peptide)** | | |  |  |  |  |  |  |
| Variable | *DCS* |  | *GoDARTS* |  | *ANDIS* |  | *ACCELERATE* |  |
|  | *HR* | *P-value* | *HR* | *P-value* | *HR* | *P-value* | *HR* | *P-value* |
| Age | 0.98[0.97-0.99] | <0.0001 | 0.97[0.96-0.98] | <0.0001 | 1.00[0.99-1.01] | 0.509 | 0.97[0.96-0.99] | 0.01 |
| Sex | 0.83[0.68-1.02] | 0.08 | 0.98[0.87-1.11] | 0.7572 | 1.14[0.97-1.34] | 0.1034 | 1.13[0.77-1.67] | 0.53 |
| BMI | 1.02[1.00-1.04] | 0.02 | 1.01[1.00-1.02] | 0.0614 | 0.99[0.97-1.00] | 0.0981 | 1.00[0.98-1.03] | 0.86 |
| HDL | 0.27[0.18-0.40] | <0.0001 | 0.67[0.55-0.82] | <0.0001 | 0.64[0.50-0.81] | 0.0003 | 0.78[0.43-1.43] | 0.43 |
| C-peptide | 0.35[0.30-0.40] | <0.0001 | 1.02[0.90-1.16] | 0.7643 | 0.61[0.50-0.74] | <.0001 | 1.37[0.92-2.04] | 0.12 |
| **Model 3 (M2 + diabetes duration + glucose-lowering drugs)** | | |  |  |  |  |  |  |
| Variable | *DCS* |  | *GoDARTS* |  | *ANDIS* |  | *ACCELERATE* |  |
|  | *HR* | *P-value* | *HR* | *P-value* | *HR* | *P-value* | *HR* | *P-value* |
| Age | 0.98[0.97-0.99] | 0.0002 | 0.97[0.96-0.97] | <0.0001 | 1.00[0.99-1.01] | 0.6339 | 0.96[0.94-0.98] | <0.0001 |
| Sex | 0.81[0.66-1] | 0.05 | 0.99[0.88-1.12] | 0.9284 | 1.13[0.97-1.33] | 0.1295 | 1.03[0.70-1.53] | 0.88 |
| BMI | 1.02[1-1.04] | 0.02 | 1.01[0.99-1.02] | 0.1121 | 0.99[0.97-1.01] | 0.1311 | 1.01[0.98-1.04] | 0.57 |
| HDL | 0.29[0.2-0.43] | <0.0001 | 0.59[0.49-0.72] | <0.0001 | 0.70[0.55-0.89] | 0.0035 | 0.80[0.44-1.45] | 0.46 |
| C-peptide | 0.36[0.31-0.42] | <0.0001 | 0.96[0.85-1.09] | 0.5516 | 0.67[0.55-0.82] | <.0001 | 1.50[1.02-2.22] | 0.04 |
| Diabetes duration | 1.00[1.00-1.00] | 0.39 | 1.06[1.03-1.08] | <0.0001 | 0.84[0.61-1.17] | 0.3099 | 1.05[1.03-1.07] | <0.0001 |
| Glucose lowering drugs | 3.42[2.63-4.45] | <0.0001 | 2.33[1.99-2.73] | <0.0001 | 0.52[0.45-0.61] | <.0001 | 4.73[1.50-14.92] | <0.0001 |

*M1, model 1; M2, model 2; HR, hazard ratio. Numbers between brackets represent confidence intervals*
